# Epigenome-wide Association Analysis of Mitochondrial Heteroplasmy Provides Insight into Molecular Mechanisms of Disease

**DOI:** 10.1101/2024.12.05.24318557

**Authors:** Meng Lai, Kyeezu Kim, Yinan Zheng, Christina A. Castellani, Scott M. Ratliff, Mengyao Wang, Xue Liu, Jeffrey Haessler, Tianxiao Huan, Lawrence F. Bielak, Wei Zhao, Roby Joehanes, Jiantao Ma, Xiuqing Guo, JoAnn E. Manson, Megan L. Grove, Jan Bressler, Kent D. Taylor, Tuuli Lappalainen, Silva Kasela, Thomas W. Blackwell, Nicole J. Lake, Jessica D. Faul, Kendra R. Ferrier, Lifang Hou, Charles Kooperberg, Alexander P. Reiner, Kai Zhang, Patricia A. Peyser, Myriam Fornage, Eric Boerwinkle, Laura M. Raffield, April P. Carson, Stephen S. Rich, Yongmei Liu, Daniel Levy, Jerome I. Rotter, Jennifer A. Smith, Dan E. Arking, Chunyu Liu, NHLBI Trans-Omics for Precision Medicine (TOPMed) Consortium

## Abstract

The relationship between mitochondrial DNA (mtDNA) heteroplasmy and nuclear DNA (nDNA) methylation (CpGs) remains to be studied. We conducted an epigenome-wide association analysis of heteroplasmy burden scores across 10,986 participants (mean age 77, 63% women, and 54% non-White races/ethnicities) from seven population-based observational cohorts. We identified 412 CpGs (FDR p < 0.05) associated with mtDNA heteroplasmy. Higher levels of heteroplasmy burden were associated with lower nDNA methylation levels at most significant CpGs. Functional inference analyses of genes annotated to heteroplasmy-associated CpGs emphasized mitochondrial functions and showed enrichment in cardiometabolic conditions and traits. We developed CpG-scores based on heteroplasmy-count associated CpGs (MHC-CpG scores) using elastic net Cox regression in a training cohort. A one-unit higher level of the standardized MHC-CpG scores were associated with 1.26-fold higher hazard of all-cause mortality (95% CI: 1.14, 1.39) and 1.09-fold higher hazard of CVD (95% CI: 1.01–1.17) in the meta-analysis of testing cohorts, adjusting for age, sex, and smoking. These findings shed light on the relationship between mtDNA heteroplasmy and DNA methylation, and the role of heteroplasmy-associated CpGs as biomarkers in predicting all-cause mortality and cardiovascular disease.

## INTRODUCTION

In addition to producing molecular energy, mitochondria are central to biochemical processes including metabolite synthesis, ion homeostasis, oxidative stress, and apoptosis.^1–3^ Defects in mitochondria are implicated in various human diseases including neurodegenerative disease,^4^ cancer,^5^ and cardiovascular disease (CVD).^6,7^ Mitochondria have their own genome, a double-stranded DNA molecule (mtDNA), which is 16.6 kb in size and exists in multiple copies per cell.^7^ This gives rise to two important characteristics: mtDNA copy number (mtDNA CN), a measure of the average number of mtDNA molecules per cell, and heteroplasmy, whereby different mtDNA alleles coexist in the same sample.^7^

DNA methylation, the most commonly studied form of epigenetic modification, has been linked to mtDNA variation as a mechanism through which mtDNA may influence the etiology of complex diseases by regulating gene expression.^8,9^ mtDNA-directed changes in DNA methylation are one of the mechanisms by which mtDNA and nuclear DNA (nDNA) cross-talk. A recent study found that global nDNA methylation levels vary in human cybrid cells, in which the cells contained the same nuclear DNA but different mtDNA haplogroups.^10^ Another study showed that mammary tumor metastasis was altered through changes in DNA methylation levels in a hybrid mouse model with nDNA from one strain and mtDNA from another strain.^11^ The subsequent gene expression levels were also altered in response to the change in nDNA methylation levels in both studies.^10,11^ Recent epigenome-wide association studies (EWAS) and meta-analyses identified CpG sites associated with mtDNA CN.^12,13^ Several of these CpG sites were associated with mortality and CVD, which suggests that mtDNA CN may play a role in adverse health outcomes via specific nDNA methylation markers.^12,13^

Despite evidence of the interplay between mtDNA and nDNA methylation, knowledge gaps remain, particularly regarding the link between mtDNA heteroplasmy and nDNA methylation. Coupled with the prior evidence, we hypothesized that mtDNA heteroplasmy is associated with methylation levels of nDNA, which may play a role in age-related health outcomes such as CVD and mortality. Leveraging data from multiple large-scale cohort studies incorporating diverse race groups (**Figure 1**), we aimed to identify nDNA CpG sites that are associated with mtDNA heteroplasmy and to investigate whether the heteroplasmy-associated methylation markers are associated with all-cause mortality and CVD.

**Figure 1.**
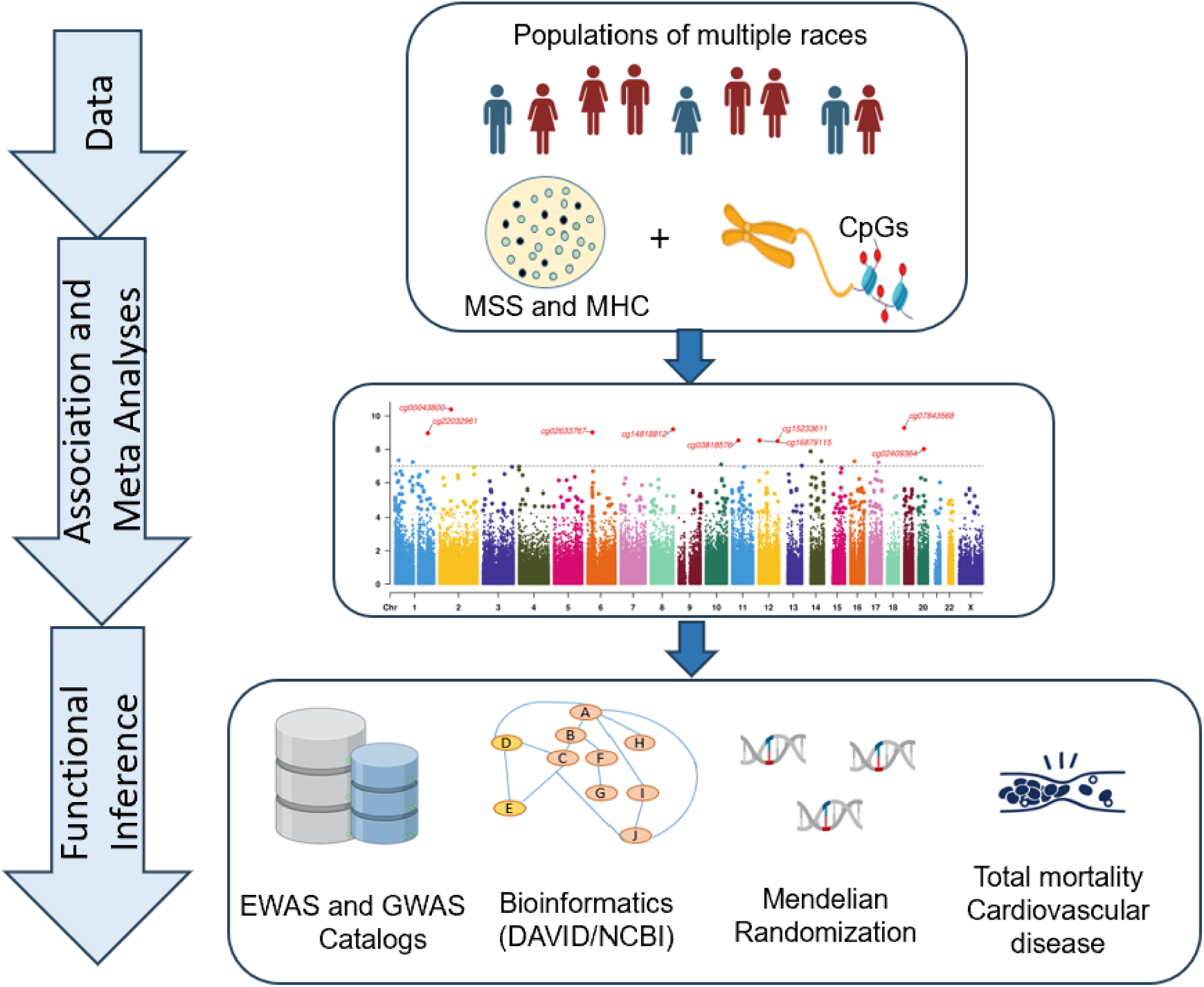
Study flowchart. Whole genome sequencing was used to identify mtDNA heteroplasmy in 10,964 participants from seven cohorts. Cohort- and race-specific analyses were conducted to investigate the association of mtDNA heteroplasmy and nuclear DNA (nDNA) methylation levels. Meta-analyses were performed in both pooled and race-specific samples. Significant CpGs identified from the meta-analyses (FDR<0.05) underwent further investigation through functional analyses. DNA methylation scores were constructed using heteroplasmy-associated CpGs and used to conduct the association analysis with total mortality.

## RESULTS

### Participant characteristics

The study participants (n=10,986, 54% non-White participants, 63% women) were mostly middle-aged (mean age 57) participants across the seven cohorts (**Supplemental Tables 1 - 2**). The Women’s Health Initiative (WHI) included only women. All the other cohorts consisted of more women than men, ranging from 50% to 72% women. A small proportion of participants were current smokers, ranging from 8% to 22%. Former smokers ranged from 15% to 45% and never smokers ranged from 41% to 65%. The predicted smoking score and reported smoking status were consistent for most participants (**Supplemental Figure 1**). Approximately one-third of the participants carried at least one heteroplasmic variant. About 8.3% to 18.8% of participants exhibited an MSS between 0.01 and 0.25, 2.7% to 5.8% displayed an MSS between 0.251 and 0.5, and 5.4% to 10.8% had an MSS greater than 0.5 (**Supplemental Tables 1 - 2**).

### Association and meta-analyses of heteroplasmy with DNA methylation

We performed cohort- and race-specific association analyses between mtDNA heteroplasmy (MHC and MSS) and DNA methylation levels using linear or linear mixed regression models, followed by meta-analyses (n=10,986) (**Figure 2A**). As main results, we report MSS- and MHC-associated CpG sites (FDR *p* < 0.05) present on both the Infinium HumanMethylation450 and Illumina MethylationEPIC arrays (**Supplemental Table 3**). The CpG sites identified solely in the Illumina MethylationEPIC BeadChip array (n=5581 participants, *p*<1e-4) are included in the Supplementary Materials. Race-specific meta-analysis revealed consistent directionality for most CpGs across both White and Black participants (**Supplemental Figures 3-6**). Below we report results from pooled analyses.

**Figure 2.**
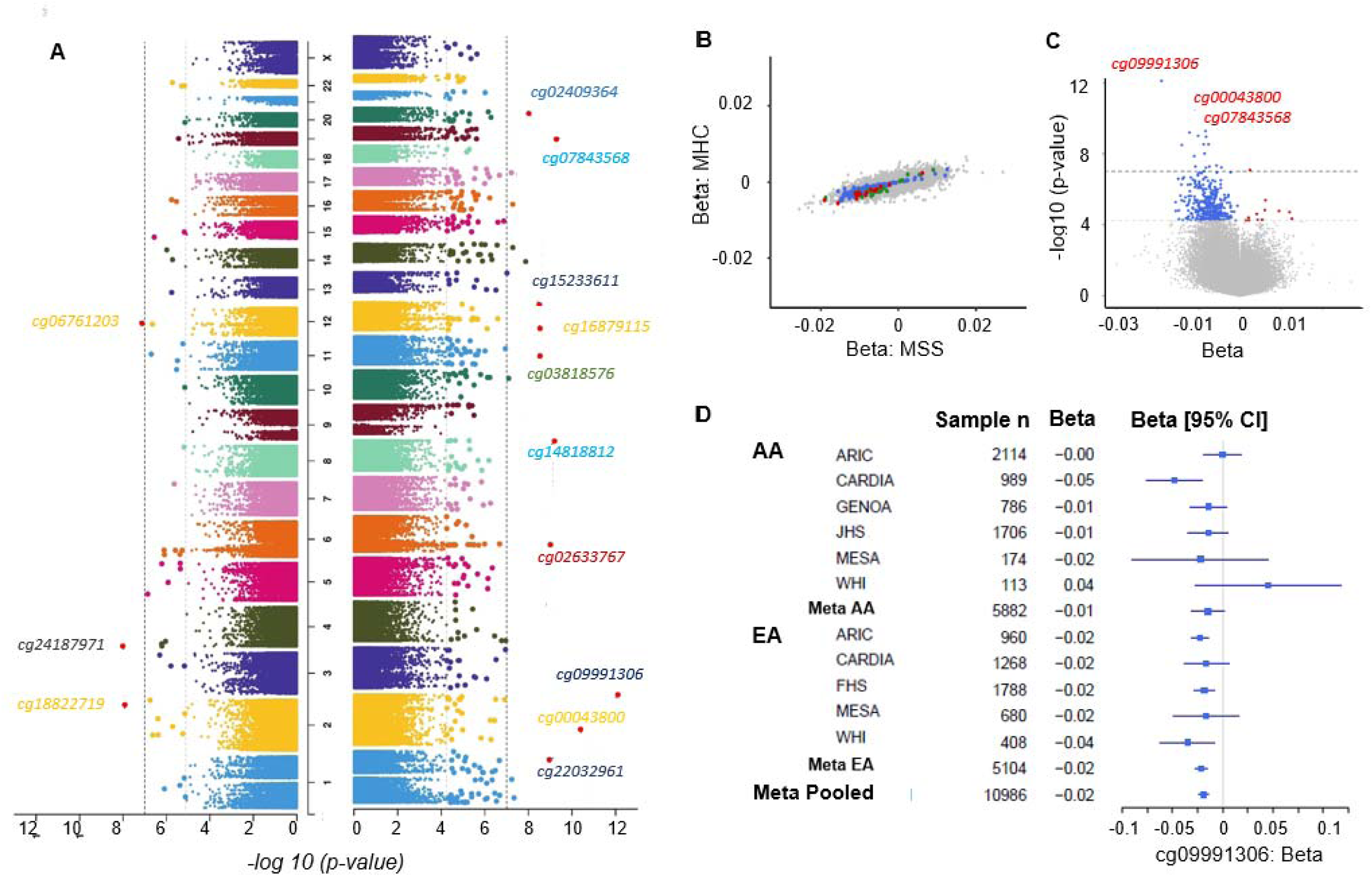
Association and meta-analyses of mtDNA heteroplasmy with DNA methylation. A. Manhattan plot: Cohort- and race-specific association analyses were conducted, followed by meta-analyses in pooled samples (n=10,964) to identify common CpGs associated with heteroplasmy across ancestries. A total of 51 CpGs displayed associations with MHC (left) and 380 CpGs displayed associations with MSS (right subplot) at FDR-adjusted p-value < 0.05. The grey dotted line is the p-value corresponding to FDR=0.05 cutoff, the black dotted line is the more stringent Bonferroni-corrected p-value cutoff 1e-7. B. Comparison of effect sizes (beta values) of the genome-wide associations and meta-analysis of DNAm with MSS to MHC. The red dots represent the CpGs significantly associated with both MSS and MHC (FDR<0.05). The blue dots are the CpGs found significantly associated with MSS (FDR<0.05) but not MHC. The green dots are the CpGs found significantly associated with MHC (FDR<0.05) but not MSS. The effect sizes are highly correlated (Pearson r =0.71). C. Volcano plot of association and meta-analysis of DNA methylation with MSS: The majority of significant CpGs (96.8% with FDR<0.05) identified in MSS analyses displayed negative associations with heteroplasmy. The blue points on the plot are the CpGs (FDR<0.05) with negative effect size, and the red points are the CpGs (FDR<0.05) with positive effect size. D. Forest plot for top CpG cg09991306 in MSS: All studies except for WHI AA shows consistent direction of effect. FHS, Framingham Heart Study; JHS, Jackson Heart Study; HRS, Health and Retirement Study; MESA, Multi-Ethnic Study of Atherosclerosis Study, WHI, Women’s Health Initiative. ARIC, Atherosclerosis Risk in Communities; CARDIA, Coronary Artery Risk Development in Young Adults; FHS, Framingham Heart Study; GENOA, Genetic Epidemiology Network of Arteriopathy; JHS, Jackson Heart Study; MESA, Multi-Ethnic Study of Atherosclerosis Study, WHI, Women’s Health Initiative

We observed an appropriate genomic control (λ_GC_) with MSS (λ_GC_ = 0.94) and a conservative λ_GC_ with MHC (λ_GC_ =0.83) (**Supplementary Table 4**). We observed consistent effect sizes for the genome-wide associations of DNA methylation with MSS and MHC, with a Pearson correlation coefficient of 0.71. This reflects the moderately high correlation between the two heteroplasmy metrics across the cohorts (**Figure 2B, Supplemental Table 5**). Using Bonferroni correction (*p* < 1e-7), we identified 3 significant CpGs from the MHC analysis and 18 CpGs from the MSS analysis. At FDR *p* < 0.05, we identified 412 unique CpGs associated with MSS (n=380) and MHC (n=51); these identified CpGs are distributed across the genome (**Figure 2A, Table 1, Supplemental Tables 6 & 7**). The majority of CpGs (84.3% for MHC and 96.8% for MSS with FDR *p* < 0.05) displayed negative associations with heteroplasmy, meaning that a higher level of heteroplasmy was associated with a lower level of DNA methylation (**Figure 2C**). The directionality of the association was mostly consistent across the cohorts for significant CpGs as exampled in **Figure 2D**.

**Table 1.**
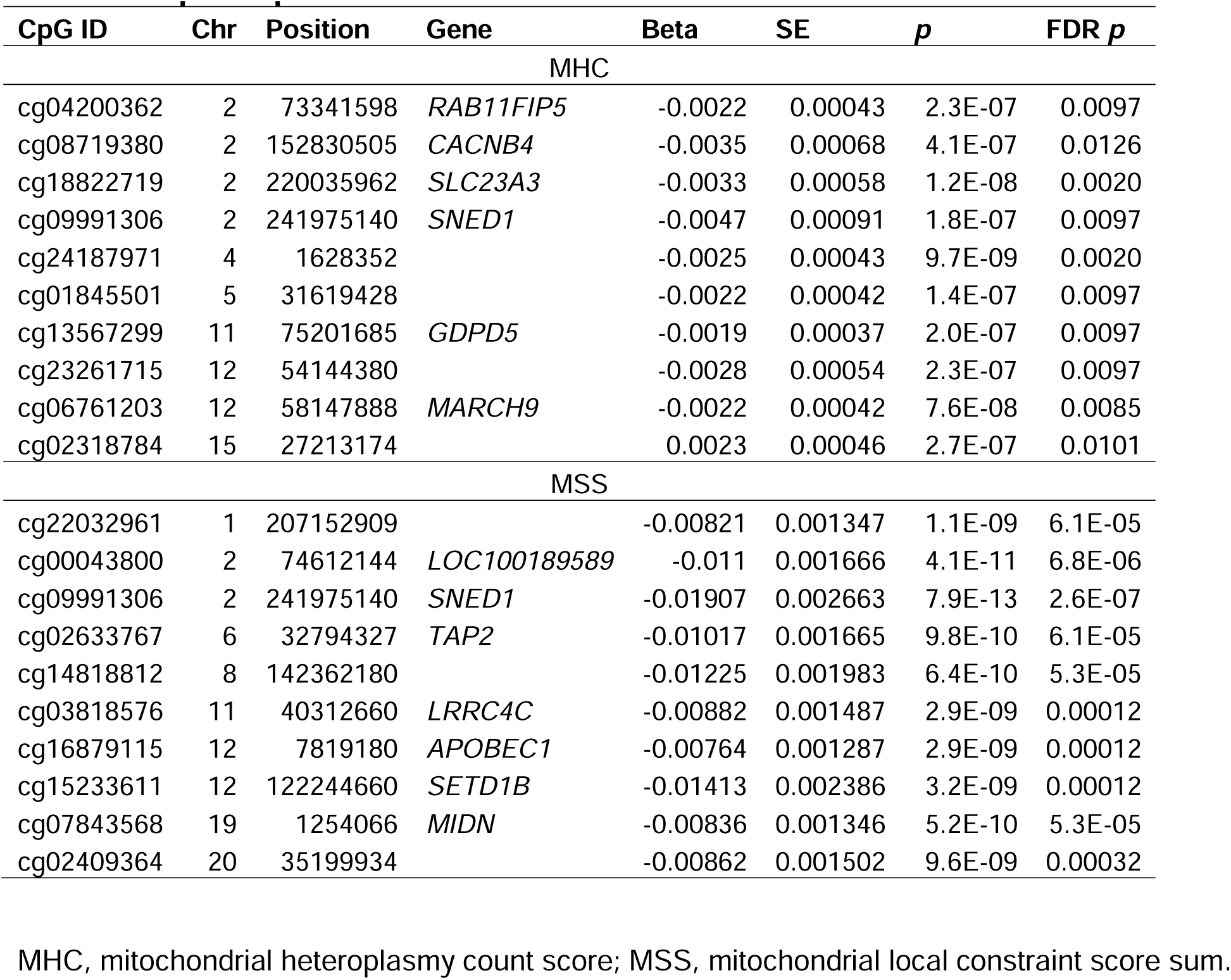
Top 10 CpGs associated with MHC and MSS.

| CpG ID | Chr | Position | Gene | Beta | SE | *p* | FDR *p* |
| --- | --- | --- | --- | --- | --- | --- | --- |
| MHC |  |  |  |  |  |  |  |
| cg04200362 | 2 | 73341598 | *RAB11FIP5* | -0.0022 | 0.00043 | 2.3E-07 | 0.0097 |
| cg08719380 | 2 | 152830505 | *CACNB4* | -0.0035 | 0.00068 | 4.1E-07 | 0.0126 |
| cg18822719 | 2 | 220035962 | *SLC23A3* | -0.0033 | 0.00058 | 1.2E-08 | 0.0020 |
| cg09991306 | 2 | 241975140 | *SNED1* | -0.0047 | 0.00091 | 1.8E-07 | 0.0097 |
| cg24187971 | 4 | 1628352 |  | -0.0025 | 0.00043 | 9.7E-09 | 0.0020 |
| cg01845501 | 5 | 31619428 |  | -0.0022 | 0.00042 | 1.4E-07 | 0.0097 |
| cg13567299 | 11 | 75201685 | *GDPD5* | -0.0019 | 0.00037 | 2.0E-07 | 0.0097 |
| cg23261715 | 12 | 54144380 |  | -0.0028 | 0.00054 | 2.3E-07 | 0.0097 |
| cg06761203 | 12 | 58147888 | *MARCH9* | -0.0022 | 0.00042 | 7.6E-08 | 0.0085 |
| cg02318784 | 15 | 27213174 |  | 0.0023 | 0.00046 | 2.7E-07 | 0.0101 |
| MSS |  |  |  |  |  |  |  |
| cg22032961 | 1 | 207152909 |  | -0.00821 | 0.001347 | 1.1E-09 | 6.1E-05 |
| cg00043800 | 2 | 74612144 | *LOC100189589* | -0.011 | 0.001666 | 4.1E-11 | 6.8E-06 |
| cg09991306 | 2 | 241975140 | *SNED1* | -0.01907 | 0.002663 | 7.9E-13 | 2.6E-07 |
| cg02633767 | 6 | 32794327 | *TAP2* | -0.01017 | 0.001665 | 9.8E-10 | 6.1E-05 |
| cg14818812 | 8 | 142362180 |  | -0.01225 | 0.001983 | 6.4E-10 | 5.3E-05 |
| cg03818576 | 11 | 40312660 | *LRRC4C* | -0.00882 | 0.001487 | 2.9E-09 | 0.00012 |
| cg16879115 | 12 | 7819180 | *APOBEC1* | -0.00764 | 0.001287 | 2.9E-09 | 0.00012 |
| cg15233611 | 12 | 122244660 | *SETD1B* | -0.01413 | 0.002386 | 3.2E-09 | 0.00012 |
| cg07843568 | 19 | 1254066 | *MIDN* | -0.00836 | 0.001346 | 5.2E-10 | 5.3E-05 |
| cg02409364 | 20 | 35199934 |  | -0.00862 | 0.001502 | 9.6E-09 | 0.00032 |

MHC, mitochondrial heteroplasmy count score; MSS, mitochondrial local constraint score sum.

Several top CpGs associated with MSS and MHC are located in genes encoding proteins involved in essential cellular functions. For example, cg09991306 (beta = - 0.019, FDR p = 2.6e-7), located in the enhancer region of *SNED1* (Sushi, Nidogen, and EGF-like domains) on chromosome 2 at locus q37.3, displayed the most significant association with MSS and was also associated with MHC (beta = −0.0047, FDR *p* = 0.01) (**Table 1**). The *SNED1* gene codes for an extracellular matrix (ECM) protein present in many tissues.^14^ Another top CpG, cg02633767 (beta = −0.010, FDR *p*= 6.1E-5 with MSS; beta=-0.0028, FDR *p* = 0.015 with MHC), is located in transporter 2, ATP binding cassette subfamily B (*TAP2*)^15^ on chromosome 6. The TAP2 transporter is associated with antigen processing and plays a crucial role in the immune system.^16^

### Functional inference of heteroplasmy-associated CpGs

### Heteroplasmy-associated CpGs and their associated traits

We queried the MRC-IEU EWAS Catalog^17^ to link previously reported diseases/traits with the 412 CpGs associated (FDR *p* < 0.05) with MSS and MHC. We identified 32 CpG-trait associations (27 unique CpGs associated with six traits [*p* < 1e-7] in studies with more than 5000 participants in the MRC-IEU EWAS Catalog ^17^ (**Supplemental Table 8**). The six traits included alcohol consumption per day (associated with 4 CpGs), body mass index (association with 5 CpGs), C-reactive protein (associated with 2 CpGs), educational attainment (associated with 1 CpG), prenatal smoke exposure (associated with 5 CpGs), and smoking (associated with 15 CpGs) (**Figure 3A**).

**Figure 3.**
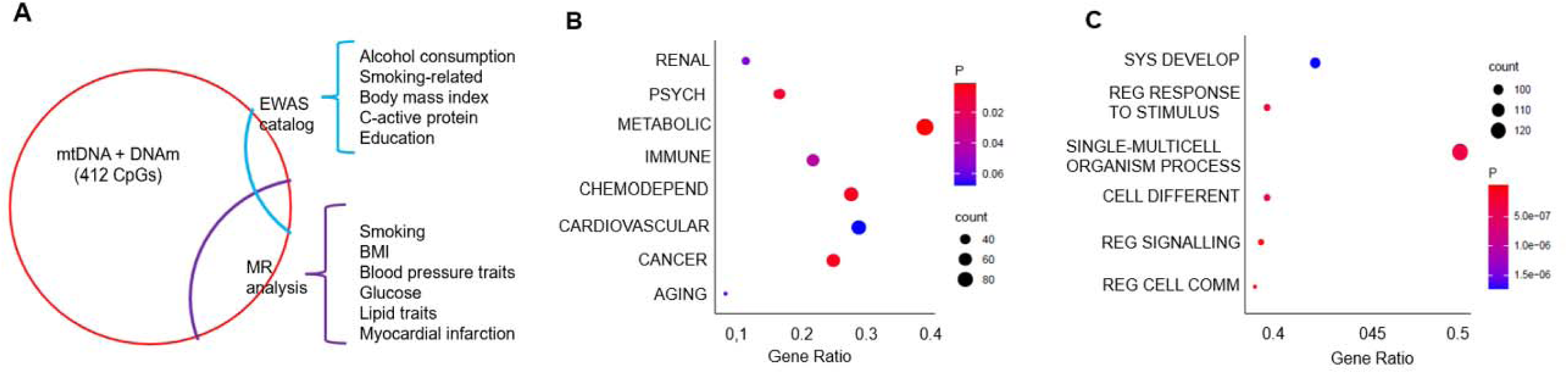
Functional Analyses. A. Out of the 412 heteroplasmy-associated CpGs, 27 CpGs are associated with 6 traits in the EWAS catalog, including alcohol consumption, BMI, C-reactive protein, educational attainment, and smoking-related traits. MR analysis inferred causal relationship between 45 CpGs and cardiovascular disease risk factors including myocardial infarction, BMI, systolic blood pressure, diastolic blood pressure, hypertension, fasting glucose, cholesterol, HDL cholesterol, LDL cholesterol, and triglycerides. B, C. Enrichment analysis using disease pathways (DAVID) show the annotated genes are enriched in metabolic traits and multicellular organism process (e.g. cell interaction).

### Transcriptomic implication of CpGs associated with heteroplasmy

To investigate possible downstream consequences of DNA methylation, we retrieved the gene transcripts associated with the identified CpGs (eQTM) from the Framingham Heart Study eQTM database.^18^ We found 49 heteroplasmy-associated CpGs exhibiting significant associations with the transcripts of 99 *cis*-genes within 1 Mb of a CpG (*p* < 1e-7) (**Supplemental Table 9**). Of the 49 CpGs, 15 displayed significant associations with the transcript levels of the genes where the CpGs were situated. For example, cg02633767, located in the 3’-untranslated region of *TAP2* (chromosome 6) was significantly associated with expression of *TAP2* (p=10e-29). Seventeen CpGs were associated with transcript levels of multiple genes within 1 Mb of a CpG. cg00589850 was associated with 22 protocadherin (*Pcdh*) genes clustered in a single genomic locus on chromosome 5 (q31.3).^19^ cg00589850 is located in the intronic regions of the *Pcdh* genes, explaining 2%-10% of the variance in the transcript levels of these genes. Eleven CpGs were significantly associated with long non-coding RNA (lncRNA) transcript levels. For example, cg17394978 in *IRF1* (Interferon regulatory factor 1) was associated with AC116366.1, a lncRNA that plays a critical role in tumor immunity (**Supplemental Table 9**).^20,21^

### Gene set enrichment analysis

A total of 255 genes were annotated to the 412 CpGs associated with MSS and MHC, and were then used for functional enrichment analysis with DAVID, a comprehensive database for functional annotation, disease association and Gene Ontology (GO) analyses.^22^ The 255 genes were significantly enriched in a broad selection of diseases such as metabolic conditions (FDR *p* = 0.007), cancer (FDR *p* = 0.015), and chemical dependency (addicted to drugs, nicotine, or alcohol) (FDR *p* = 0.015), among others (**Figure 3B, Supplemental Table 10**). GO analysis identified pathways, such as signaling (e.g., regulation of signaling [GO:0023051], FDR *p* = 1.5e-7), cell differentiation and system development (e.g., cell differentiation [GO:0030154], FDR *p* = 2.9e-7), and immune process (e.g., T cell differentiation [GO:0030217], FDR *p* = 0.017) (**Figure 3C, Supplemental Table 11**). GO analysis also identified cellular components involving synaptic functions and neurotransmission (e.g., synapse [GO:0045202], FDR *p* =0.047), and membrane components (e.g., endomembrane system [GO:0012505], FDR *p*= 0.010) (**Supplemental Table 12**).

### Genes involved in mitochondrial assembly and function

Through functional annotation with DAVID^22^ and MitoCarta3.0.^23^, we identified 27 genes known or predicted to be involved in mitochondrial activities (**Supplemental Table 13**). Among these, nine genes encode proteins constituting the mammalian mitochondrial proteome according to MitoCarta3.0,^23^ such as fragile histidine triad diadenosine triphosphatase (*FHIT*), ferritin heavy chain 1 (*FTH1*), and mitochondrial ribosomal protein L23 (*MRPL23*). Most of the remaining 18 genes encode proteins involved in activities dependent on NAD+/NADP+/NADPH or may impact the phosphorylation pathway. For instance, *CRYM* (crystallin mu) encodes a protein found in both the nucleus and mitochondria, playing a role in oxidoreductase.^24^ (**Supplemental Table 13**). *mQTLs for heteroplasmy-associated CpGs*

Investigating DNA methylation quantitative trait loci (mQTL) and their associated GWAS traits/diseases may help identify a common genetic basis underlying heteroplasmy and DNA methylation, as well as their relationship to human traits/diseases.^25,26^ We identified 110,898 mQTL-CpGs pairs (*p* < 1e-7) from 104,160 distinct SNPs and 328 heteroplasmy-associated CpGs. We linked the 9526 GWAS results (*p* < 5e-8) from 1287 traits to these mQTLs involving 216 CpGs (**Supplemental Table 14**). Traits/diseases associated with the lead mQTLs included a wide range of traits, such as CVD (e.g., coronary artery disease) and risk factors (e.g., BMI, blood pressure traits, lipids, and diabetes; smoking and alcohol consumption), and immune functions (e.g., platelet count) (**Supplemental Table 14**).

### Mendelian randomization (MR) analysis of heteroplasmy-associated CpGs for CVD risk and all-cause mortality

We performed MR analysis to investigate possible causal relations between heteroplasmy-associated CpG sites (FDR *p* < 0.05) and CVD-related traits and all-cause mortality. We identified significant *cis*-mQTLs (*p* < 5e-8) for 216 (of 412) heteroplasmy-associated CpGs (FDR *p* < 0.05). We presented the most significant results for the fourteen traits related to cardiovascular disease (FDR *p* <0.05) (**Supplemental Table 15**). For example, cg03732020 in *NR1H3* was inferred to have causal associations with body mass index (MR FDR *p* = 3.1e-42) and HDL cholesterol (MR FDR *p* = 5.5e-90). cg02639359 in FCH domain only 1 (*FCHO1*) gene showed evidence for causal effects for myocardial infarction (MR FDR *p* = 1.3e-7). In addition, cg07773769 in myocyte enhancer factor-2-activating motif and SAP domain-containing transcriptional regulator (*MAMSTR*) gene showed evidence for causal effects for myocardial infarction (MR FDR *p* = 1.3e-7).

### DNA methylation sites associated with both mtDNA CN and heteroplasmy

We previously identified DNA methylation signatures of mtDNA CN.^13^ Three CpGs (cg03732020, cg09109520, and cg02318784) associated with MSS/MHC (FDR *p* < 0.05) displayed *p* < 1e-4 (∼0.05/412) in their associations with mtDNA CN. cg03732020 is located in the nuclear receptor subfamily 1 group H member 3 (*NR1H3*) on chromosome 11, and cg09109520 is located in adhesion G protein-coupled receptor G1 (*ADGRG1* on chromosome 16) encoding G protein-coupled receptor 56 (*GPR56*). Several genes – *ARHGEF10*, *C14orf73*, *DNMT3A*, *LFNG*, *PRDM16*, *TCEA3*, and *VARS* – contain different CpGs that are associated with either heteroplasmy or mtDNA CN, indicating that these genes show varying patterns of methylation related to these two mtDNA features, exampled in *VARS* (**Supplemental Figure 7** & **Supplemental Table 16**).

### Association analysis of heteroplasmy-associated DNA methylation score with all-cause mortality

We observed 347 deaths with a median follow-up of 13 years among 3,418 participants in the FHS. In the other cohorts, we observed a total of 806 deaths with a median follow-up of 14 to 21 years across the JHS, MESA, and WHI cohorts (**Supplemental Table 17**). The HRS cohort had a median follow-up of 3 years for mortality (Supplemental Table 17).

We applied elastic net Cox regression to heteroplasmy-associated CpGs and identified 29 CpGs associated with MHC for all-cause mortality, using the FHS cohort as the training set (**Supplemental Table 18**). We constructed a weighted MHC-CpG score and found that a one standard deviation (SD) higher level of this score was associated with a 1.42-fold higher hazard of all-cause mortality (95% CI: 1.31, 1.51), adjusting for age, sex, and smoking. In the testing samples, we found that a one-SD higher level of the weighted MHC-CpG score was associated with a 1.14-fold higher hazard of all-cause mortality (95% CI: 1.04, 1.26) in the meta-analysis of JHS, MESA, and WHI cohorts, and a 1.26-fold higher hazard of all-cause mortality (95% CI: 1.14, 1.39) in the meta-analysis of JHS, MESA, WHI, and HRS cohorts, adjusting for age, sex, and smoking (**Figure 4**).

**Figure 4.**
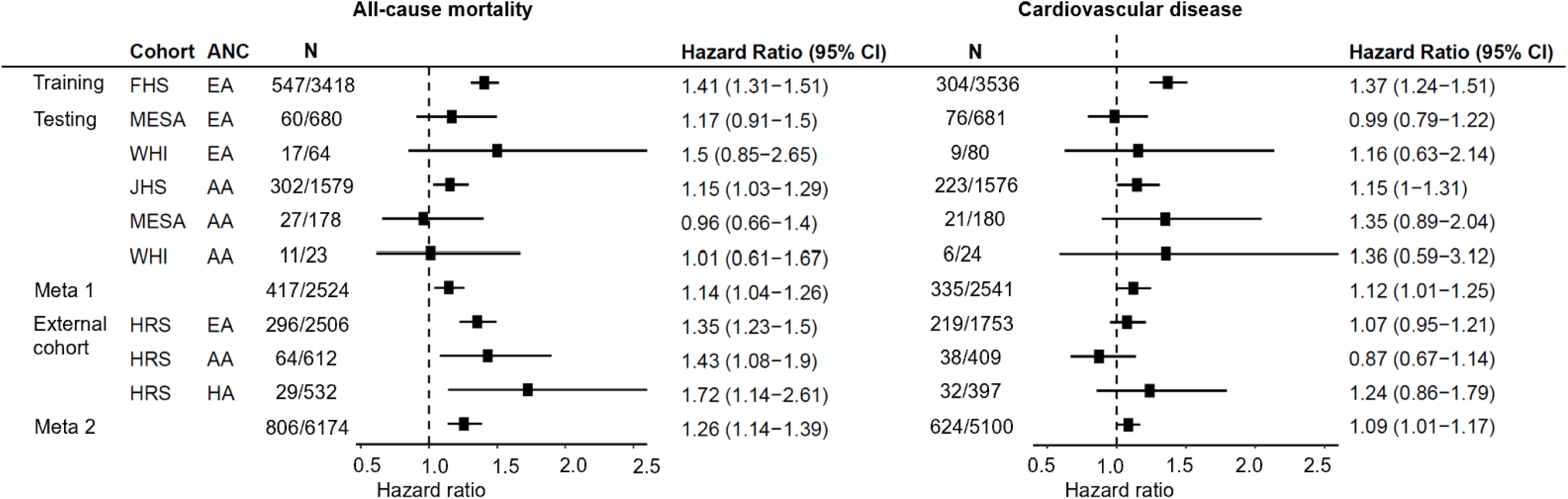
Forest plot: Association analysis of heteroplasmy-count associated DNA methylation score with mortality and incident CVD. We applied elastic net Cox regression to heteroplasmy count (MHC) associated CpGs and selected 29 CpGs for all-cause mortality and 13 CpGs for CVD using the Framingham Heart Study (FHS) as the training sample. The forest plot illustrates hazard ratios (HR) with 95% confidence intervals (CI) for association analyses of the weighted MHC-CpG scores with mortality and incident CVD across the training and testing cohorts, adjusting for age, sex, and smoking status (never, former, and current). Testing was conducted in JHS, MESA, and WHI. Testing was also conducted in HRS, which was not used for selecting heteroplasmy-associated DNA methylation. CVD, cardiovascular disease; ARIC, Atherosclerosis Risk in Communities; CARDIA, Coronary Artery Risk Development in Young Adults; FHS, Framingham Heart Study; GENOA, Genetic Epidemiology Network of Arteriopathy; JHS, Jackson Heart Study; HRS, Health and Retirement Study; MESA, Multi-Ethnic Study of Atherosclerosis Study, WHI, Women’s Health Initiative.

Similarly, we identified 56 MSS-associated CpGs for all-cause mortality using elastic net Cox regression in the FHS cohort as the training sample (**Supplemental Table 19**). We found consistent results between the MSS-CpG weighted score and all-cause mortality in the training sample, as well as in meta-analyses of the JHS, MESA, WHI, and HRS cohorts as testing samples, adjusting for age, sex, and smoking status (**Supplemental Figure 8**). Furthermore, we observed consistent results in both the base model (age and sex adjusted) and the multi-covariate adjusted model for the associations of both MHC- and MSS-weighted scores with all-cause mortality (**Supplemental Figures 8 & 9**).

### Association analysis of heteroplasmy-associated DNA methylation score with CVD

In the FHS, 304 participants developed CVD during a median follow-up of 8 years among 3,536 participants. We observed a total of 624 incident CVD cases with a median follow-up of 14 to 16 years across the JHS, MESA, and WHI. However, HRS had a much shorter median follow-up (4 years) compared to other cohorts (**Supplemental Table 17**).

We selected 13 CpGs for MHC using elastic net Cox regression (**Supplemental Table 20**) for CVD. In the FHS testing sample, we found that a SD higher level of the weighted MHC-CpG score was associated with a 1.37-fold higher hazard of CVD (95% CI: 1.24–1.51), adjusting for age, sex, and smoking. In the testing cohorts, we found that a one-SD higher MSS-CpG score was associated with a 1.12-fold higher hazard of CVD (95% CI: 1.01–1.25) in the meta-analysis of JHS, MESA, and WHI, and a 1.09-fold higher hazard of CVD (95% CI: 1.01–1.17) in the meta-analysis of JHS, MESA, WHI, and HRS (**Figure 4**).

We identified 9 MSS-associated CpGs for CVD using elastic net Cox regression in FHS (**Supplemental Table 21**). Compared to that of the weighted MHC-CpG score, the weighted MSS-CpG score showed slightly weaker associations with CVD in the training cohort and meta-analysis of JHS, MESA, WHI, and HRS as the testing samples in both base model and the model with smoking as an additional covariate (**Supplemental Figure 10**). Adjusting for additional multi-covariates further attenuated the associations for both weighted MHC- and MSS-scores (**Supplemental Figure 11**).

## DISCUSSION

We examined the associations between nuclear DNA (nDNA) methylation and mitochondrial DNA (mtDNA) heteroplasmy in 10,964 individuals from seven population-based cohorts, primarily middle-aged individuals from diverse racial and ethnic backgrounds. Our analysis identified 412 unique CpGs with differential methylation linked to mtDNA heteroplasmy, with a greater number of CpGs associated with mitochondrial stress signal (MSS) than with mitochondrial health condition (MHC). Higher heteroplasmy burden was consistently linked to lower nDNA methylation levels. Functional analyses of genes annotated to heteroplasmy-associated CpGs highlighted mitochondrial functions and revealed enrichment in cardiometabolic traits. We developed CpG scores based on these CpGs and found that higher scores were associated with increased risks of total mortality and cardiovascular disease.

Our findings align with previous evidence suggesting cross-talk between nDNA and mtDNA.^27–29^ This interaction involves bidirectional information flow and coordination in epigenetic processes that regulate a cell’s resp.onse to external cues.^29^ This is supported by our analysis of significant CpG-annotated genes, which are implicated in signaling, cell differentiation, and system development. For instance, *SNED1* (cg09991306) encodes an extracellular matrix protein critical for dynamic protein assembly in multicellular organisms, influencing developmental processes and tissue organization.^30^ *TAP2* (cg02633767) encodes a membrane-associated protein belonging to the ATP-binding cassette (ABC) superfamily, facilitating the transport of various molecules across the plasma membrane, intracellular membranes of the endoplasmic reticulum, peroxisome, and mitochondria.^31^ Moreover, our study identified over two dozen nDNA genes associated with heteroplasmy-related CpGs (**Supplemental Table 13**) involved in mitochondrial biosynthesis, mtDNA replication, and methylation regulation. Examples include *MRPL23*, which encodes a 39S subunit protein aiding in protein synthesis within the mitochondrion,^32^ and *FTH1*, encoding ferritin heavy chain 1. FTH1 regulates multiple physiological processes, including oxidative stress, inflammation, and a type of programmed cell death characterized by altered mitochondrial morphology and increased lipid peroxides.^33–35^ *DNMT3A* encodes DNA-methyltransferase 3A, which localizes to mitochondria and maintains CpH methylation in neurons in vivo.^36^ Knockout of *DNMT3A* perturbs mtDNA regional methylation patterns, while overexpression of *DNMT3A* greatly increases mtDNA methylation and strand bias.^36^

We have identified CpGs associated with mtDNA CN in our previous study.^13^ mtDNA CN reflects the quantity of mtDNA molecules, while heteroplasmy levels represent the genetic variation within the mtDNA population in an individual. We compared the CpGs associated with two mitochondrial metrics and found few CpGs were commonly associated (p<1e-7) with both mtDNA heteroplasmy level and CN, suggesting that distinct DNA methylation patterns in the nuclear genome are related to these two mtDNA features. In contrast, certain genes harbor different CpGs associated with these two features. For example, multiple CpGs within the *VARS* gene showed significant or suggestive associations with both mtDNA heteroplasmy and CN (**Supplemental Figure 7**), suggesting that these genes might be involved in crosstalk with mtDNA in their physiological roles in humans.

Our MR analysis supports prior research on the role of DNA methylation in CVD risk. For example, methylation levels of *NR1H3* may causally influence several CVD risk factors such as blood pressure BMI, fast blood insulin, and HDL cholesterol (**Supplemental Table 15**). Previous research reported CpGs at *NR1H3* were associated with CVD risk factors like aging and smoking.^37^ *NR1H3* encodes a protein in the nuclear receptor superfamily that is important for regulating macrophage function and transcriptional programs that regulate lipid homeostasis and inflammation.^38^ Additionally, cg03732020 at *NR1H3* has been associated with mtDNA CN in our previous studies.^13^ Our results suggest that mtDNA heteroplasmic variants may influence physiological traits through methylation changes in multiple nDNA CpGs. Investigating how heteroplasmic variants may causally affect nDNA methylation levels is a key direction for future research.

We developed CpG-scores based on heteroplasmy-count associated CpGs (MHC-CpG scores) using elastic net Cox regression in a training cohort. A one-unit higher level of the standardized MHC-CpG scores were associated with 1.26-fold higher hazard of all-cause mortality (95% CI: 1.14, 1.39) and 1.09-fold higher hazard of CVD (95% CI: 1.01– 1.17) in the meta-analysis of testing cohorts, adjusting for age, sex, and smoking. These findings shed light on the relationship between mtDNA heteroplasmy and DNA methylation, and the role of heteroplasmy-associated CpGs as biomarkers in predicting all-cause mortality and cardiovascular disease.

We developed CpG scores based on MHC- and MSS-associated CpGs using elastic net Cox regression, with the FHS serving as the training cohort. We found that higher levels of MHC- and MSS-CpG scores were strongly linked to an increased hazard of all-cause mortality in the testing cohorts, with similar magnitudes. Our findings support prior research suggesting that heteroplasmy-burden scores predict all-cause mortality.^39^ Additionally, we observed that a higher MHC-CpG score was associated with an elevated hazard of CVD, though the association was notably weaker than that with all-cause mortality. Our findings suggest two potential explanations. First, nDNA methylation levels could mediate the effect of mtDNA heteroplasmy on all-cause mortality and CVD. Alternatively, our results may simply reflect an association between nDNA methylation, mtDNA heteroplasmy, and the outcome variables, without implying a causal link. To test whether nDNA methylation mediates the effects of mtDNA heteroplasmy on all-cause mortality and CVD, future studies could employ longitudinal designs with larger, more diverse cohorts. Additionally, functional studies using experimental models (e.g., cell lines or animal models) could manipulate mtDNA heteroplasmy to evaluate its impact on nDNA methylation and associated outcomes Despite using consistent methods to identify heteroplasmy, we observed differences in cohort- and race-specific association analyses. This variability is likely due to genetic differences, unaccounted social and environmental factors, and variations in the conditions under which an experiment is conducted. To address this, we employed a random effects model in our meta-analysis. While studying DNA methylation and heteroplasmy in whole blood may not fully capture all tissues and cell types, blood samples are easily accessible and provide a broad overview of various tissues and organs. The study’s strength lies in its rigorous statistical analysis, employing comprehensive methods to explore the complex relationship between mtDNA heteroplasmy and DNA methylation across cohorts with diverse populations, and using a series of models to adjust for potential confounders. This approach enhances the credibility and generalizability of our findings.

In conclusion, our study identified associations between mtDNA heteroplasmy and DNA methylation. We uncovered numerous heteroplasmy-associated CpGs implicated with metabolic traits, signaling pathways, system development, and total mortality. These findings shed light on the intricate interplay between mtDNA heteroplasmy and DNA methylation, providing insights into their roles in human health and disease.

## METHODS

### Study participants

This study included 10,964 participants (mean age 57 years, women 63%, 54% non-White participants) from seven cohorts: ARIC (Atherosclerosis Risk in Communities),^40^ CARDIA (Coronary Artery Risk Development in Young Adults),^41^ FHS (Framingham Heart Study),^42,43^ GENOA (Genetic Epidemiology Network of Arteriopathy),^44^ JHS (Jackson Heart Study),^45^ MESA (Multi-Ethnic Study of Atherosclerosis),^46^ and WHI (Women’s Health Initiative).^47^ Based on prior research,^48^ most FHS participants showed high genetic similarity to European ancestry reference panels, while most JHS and GENOA participants were similar to African ancestry reference panels (mean ∼80%). Other cohorts included individuals from both self-identified groups. For simplicity, we refer to these groups as White participants and Black participants in this paper. Our analyses were stratified by self-reported race/ethnicity, without excluding genetic ancestry outliers. Thus, race/ethnicity as used in this study should not be considered equivalent to ancestry proportions. In addition, MESA included 33% Hispanic participants (**Supplemental Table 1**). Details on each cohort are described in the **Supplementary Methods.**

### Profiling and quality control of DNA methylation in whole blood

Peripheral whole blood samples from fasting blood tests were used for genomic DNA extraction and bisulfite conversion, followed by methylation profiling per the manufacturer’s protocol (Illumina Inc., San Diego, CA). Two platforms were used for DNA methylation measurement across cohorts (**Supplemental Table 1**): the Infinium HumanMethylation 450k BeadChip array (covering over 480,000 CpG sites, Illumina Inc., San Diego, CA) for FHS, WHI, and ARIC, and the Infinium MethylationEPIC BeadChip array (covering over 850,000 CpG sites, Illumina Inc., San Diego, CA) for CARDIA, GENOA, JHS, and MESA. Over 90% of CpGs from the 450k array are covered by the EPIC array. Analyses included all CpGs covered by both arrays, with subsequent analyses focusing on overlapping CpGs. To ensure data quality, we excluded CpGs with high missing rates (>20%), non-significant detection p-values (>0.01), those that were SNPs or had an underlying SNP (minor allele frequency > 5%) in participants from the 1000 Genomes Project data within 10 bp of the probes, or those mapped to multiple locations. Samples with high missing rates (>1%), poor genotype matching, or identified as outliers in clustering analysis were also excluded.^49^ Additional cohort-specific methods are detailed in the **Supplementary Methods**.

### Whole genome sequencing in whole blood

The whole blood-derived DNA in each cohort underwent whole genome sequencing (WGS) at several TOPMed contract sequencing centers.^48^ The Human Genome Sequencing Center at the Baylor College of Medicine and the Broad Institute performed WGS for ARIC and CARDIA samples. Whole genome sequencing of the FHS, WHI, and MESA samples was conducted by the Broad Institute of MIT and Harvard. Samples from JHS were sequenced at the University of Washington. WGS of GENOA was performed at the University of Washington and the Broad Institute. All the sequencing centers employed consistent data processing and sequencing processing criteria. Subsequent DNA sequence alignment of the reads to human genome build GRCh38 was also carried out at these locations. The generated BAM files were sent to TOPMed’s Informatics Research Center (IRC). For the purpose of consistency, the IRC administered re-alignment and the remake of the BAM files using a common pipeline.^48^ This study used the WGS data from Freeze 8.

### Identification and quantification of mtDNA heteroplasmy

The MToolBox software package^50^ was applied to identify heteroplasmy in mtDNA sequence reads for all cohorts except ARIC, where the mitochondrial high-performance call (mitoHPC)^51^ pipeline was applied. MToolBox removed nuclear mitochondrial DNA segments (NumtS) by remapping reads onto the reference nuclear genome (GRCh37/hg19) and applied scripts to detect nucleotide mismatches and detect indels^50^. mitoHPC is an automated pipeline to analyze mtDNA sequence reads with a circularized mitochondrial chromosome. mitoHPC extracts NumtS to build up mtDNA read sinks. mtDNA reads were further remapped to a circularized mitochondrial chromosome (chrM) to recover low coverage areas.^51^

For this study, we only considered heteroplasmic sites from single nucleotide variants (SNVs). At each mtDNA site, a variant allele was identified by comparing mtDNA sequence reads to the revised Cambridge Reference Sequence (rCRS)^52^. A variant allele fraction (VAF) was defined as the ratio of variant allele reads to the overall sequence reads observed at that mtDNA site. To minimize false positives, a heteroplasmic variant was determined using the 5%-95% threshold of VAF based on a previous study.^39^ For a mtDNA site *j* of individual *i*, the heteroplasmic variant, denoted as H_ij_, was defined by the following indicator function. If the VAF at a mtDNA site exceeded the lower or upper bound, the indicator function was assigned a value of 0:

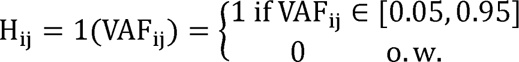

To investigate the association between heteroplasmy and DNA methylation, we constructed two continuous variables to quantify heteroplasmic burden: mitochondrial heteroplasmy count (MHC) and the mitochondrial local constraint score sum (MSS).^39,53^ The MHC of participant *i* was defined as the sum of the number of mtDNA heteroplasmic sites: MHC_i_ = ∑_j_ H_ij_. The MSS was based on the measure of the mitochondrial local constraint (MLC) score that functionally characterizes a mtDNA allele. Each mtDNA allele is assigned a MLC score between 0 and 1, and a higher MLC score indicates more harmful biological consequences.^54^ The MSS_i_ was defined as the sum of the MLC scores of variant alleles at all heteroplasmy sites in individual *i* or MSS_i_= ∑_j_m_j_H_ij_. Thus, the MSS quantifies the potential functional influence of the heteroplasmic burden for an individual. In the primary analysis, the MSS served as the main predictor in the association analyses with DNA methylation sites.

### Association and meta-analyses of mtDNA heteroplasmy burden with DNA methylation

#### Association analyses

We employed either a linear (for unrelated data) or a linear mixed (for family data) regression model to quantify the association between a mtDNA heteroplasmy burden variable and the levels of a DNA methylation site (i.e., CpG) (**Figure 1**). Our analysis framework is as follows. In the first step, the nDNA methylation residuals were calculated by regressing the nDNA methylation values of a CpG on age, age squared, sex, smoking score, white blood cell counts, and batch variants (chip IDs and rows/columns). The smoking score was calculated from 183 CpGs using the EpiSmokEr R package^55^ to provide a more objective assessment of a person’s smoking status and better reflect smoking history compared to self-report (e.g., recall bias, second-hand smoking or missing data due to reluctance to report). The methylation residuals were then modeled as the outcome variable with the mtDNA heteroplasmy burden as the explanatory variable, adjusting for age, age squared, sex, smoking score, and the year of blood draw (representing the batch variable for mtDNA measurement). ^55^ Linear mixed effect models were used to account for the random effects indicated by maternal lineage IDs in cohorts with family structures (i.e., FHS, GENOA, and JHS). To minimize the confounding effect of smoking,^39^ we explored different smoking variables in the regression analyses of methylation residuals with heteroplasmy burden, including smoking status (i.e., never, former, and current smokers), smoking score, and the combination of smoking status and smoking score. We found that the smoking score was able to capture the most smoking-related signals, and hence, we used the smoking score in our primary analysis. Analyses incorporating self-reported smoking status can be found in the supplemental materials. We conducted cohort- and race-specific association analyses. In MESA, participants of Hispanic ethnicity were combined with non-Hispanic White participants because previous findings indicate that the genetic effects tend to be similar between these racial groups.^56^ We adjusted an index variable to represent racial groups in the association analysis of the combined participants in MESA.

#### Meta-analysis

The inverse variance weighted random effects method was used to combine results across cohorts. To identify race-specific associations, we conducted separate meta-analyses for Black and White participants (**Figure 1**). To explore associations across races, we conducted a meta-analysis of all participants. We reported primary results for CpGs that were present on both the Infinium HumanMethylation450 BeadChip array and the MethylationEPIC BeadChip array in the meta-analyses. We used FDR *p* < 0.05 in meta-analysis to account for multiple testing. All subsequent analyses were conducted with these CpGs.

### Functional inference

#### Linking traits to heteroplasmy-associated CpGs

For functional inference, we mapped heteroplasmy-associated CpGs (FDR-adjusted *p* < 0.05) to disease traits by querying the MRC-IEU EWAS Catalog.^17^ We reported diseases/traits for CpGs with *p* < 1e-7 in studies with sample sizes exceeding 5,000 from the MRC-IEU EWAS Catalog.^17^

#### Gene set enrichment analysis

We used the DAVID^22^ (Database for Annotation, Visualization and Integrated Discovery) web server for functional enrichment analysis and functional annotation of genes mapped to heteroplasmy-associated CpGs (FDR *p*< 0.05). DAVID offers a robust suite of functional annotation tools aimed at decoding the biological significance of gene sets.^22^ In particular, DAVID obtains biological terms from Go Ontology and pathways from resources, such as BioCarta^57^ and Kyoto Encyclopedia of Genes and Genomes (KEGG).^58^ Statistically significant pathways were reported at FDR *p* ≤ 0.05.

#### Identification of DNA methylation quantitative trait loci (mQTL)

To explore the genetic basis of heteroplasmy-associated CpGs, we queried the mQTL database generated in the 4126 FHS participants who had both WGS and DNA methylation data^59^. The mQTL analysis identifies single nucleotide polymorphisms (SNPs) that are associated with the methylation of neighboring (*cis*-) or distant (*trans*-) CpGs. We focused on *cis*-mQTLs, i.e., SNPs residing within 1 Mb (±1 Mb) from a CpG site. Details for data generation and statistical analyses to generate the mQTL database were described previously.^59^ We examined traits that showed associations with the significant *cis*-mQTLs (i.e., SNPs) using the NHGRI-EBI GWAS Catalog.^60^ We considered SNPs that showed significant associations (*p* < 5e-8) with traits in studies with more than 5000 samples in discovery and replication analyses, or studies with more than 10,000 participants with only discovery analyses. mQTLs were used in Mendelian randomization (MR)^61^ to infer causal association between CpGs and traits related to CVD risk.

#### Identification of expression quantitative trait methylation (eQTM)

To explore possible downstream consequences of heteroplasmy-associated CpGs, we queried the eQTM database generated in the FHS.^18^ The eQTM analysis identifies CpG sites that display associations with expression of nearby (*cis*-) or remote (*trans*-) genes. In FHS, the eQTM resource was generated using DNA methylation and gene transcript levels based on RNA sequencing (i.e., RNAseq) in 2,115 study participants. Detailed QC procedures for DNA methylation and RNA sequencing have been previously described^18^. We focused on *cis*-eQTMs, which were gene transcripts whose transcription start sites were within 1 Mb of a CpG. The identified *cis*-eQTMs (i.e., nearby genes to the CpGs) were used for gene enrichment analysis (**Figure 1**)

#### Mendelian randomization (MR) analysis for CVD-related traits and all-cause mortality

To investigate whether differential methylation at heteroplasmy-associated CpGs causally influences CVD risk and mortality, we performed two-sample MR^61^ between exposures (heteroplasmy-associated CpGs) and a range of CVD and mortality-related traits as outcomes (myocardial infarction, body mass index, obesity, systolic blood pressure, diastolic blood pressure, hypertension, fasting glucose, fasting insulin, diabetes, total cholesterol, HDL cholesterol, LDL cholesterol, triglycerides, and all-cause mortality). Our in-house developed analytical pipeline, MR-Seek (https://github.com/OpenOmics/mr-seek.git), was used for MR analysis.^62^ Full summary statistics for 516 GWAS datasets were downloaded from NHGRI-EBI.

*Cis*-mQTL variants^59^ were used as the instrumental variables (IVs) in the MR analyses (**Figure 1**). We selected independent cis-mQTLs (linkage equilibrium, r^2^ < 0.001),^63^ retaining only the cis-mQTL variant with the lowest SNP-CpG p-value in each LD block. Inverse-variance weighted (IVW) MR tests were applied to combine results from multiple IVs, and used the MR-Egger method to assess horizontal pleiotropic effects.^63^ Results with a significance level of *p*<0.05 for heterogeneity were excluded. For CpG with only one IV, the Wald MR method was used to access significance. Significance levels of MR results were determined based on the Benjamini-Hochberg corrected FDR adjusted p-value with a threshold of <0.05. The most significant result was presented for each trait (outcome).

#### Examining common CpGs associated with mtDNA heteroplasmy and copy number

Previous studies have identified several CpGs associated with mtDNA CN.^12,13^ We examined the overlap between CpGs associated to mtDNA heteroplasmy and those associated with mtDNA CN. We also compared the genes annotated to both lists (i.e., CpGs associated with mtDNA heteroplasmy and those with mtDNA CN) using a relaxed threshold (*p* < 1e-4). Additionally, we investigated DNA co-methylation patterns using the coMET^64^ software to visualize patterns in selected genes containing multiple CpGs related to both mtDNA heteroplasmy and copy number.

### Association analysis of heteroplasmy-associated CpG score with all-cause mortality and cardiovascular disease (CVD)

#### Outcome definitions

We investigated whether heteroplasmy-associated CpGs were associated with all-cause mortality and CVD, given that mtDNA heteroplasmy has been associated with all-cause mortality^39^ and hypertension, a major risk factor for CVD^65^ All-cause mortality includes deaths from any cause. Incident CVD events included myocardial infarction (recognized or unrecognized or by autopsy), angina pectoris, coronary insufficiency, congestive heart failure, cerebrovascular accident, atherothrombotic infarction of brain, and death due to these conditions.

Given the correlation among many CpGs, we employed the elastic-net method with regularized Cox regression from the ‘glmnet’ R package to select heteroplasmy-associated CpGs for testing their association with all-cause mortality and CVD.

#### Selection of CpGs for predicting all-cause mortality and CVD

Given the correlation among many CpGs, we selected heteroplasmy-associated CpGs for predicting all-cause mortality and CVD using the elastic-net method with regularized Cox regression from the “glmnet” R package.^66,67^ The elastic-net method combines Ridge and Lasso penalties, allowing flexible regularization for feature selection and coefficient shrinkage despite covariate multicollinearity. We first obtained CpG residuals by regressing CpGs (i.e., those associated to MHC and MSS) on age, sex, and smoking scores. The FHS cohort was used as the training set, and we selected CpGs to predict CVD and all-cause mortality using an alpha of 0.5 and five-fold cross-validation to determine the lambda value that minimizes the mean cross-validated error.^66,67^

The selected CpGs were used to construct the heteroplasmy-associated CpG scores for predicting CVD or all-cause mortality. For individual *j*, the score *s_j_* was constructed as a weighted sum of methylation levels across heteroplasmy-associated CpG sites: *s_j_* = ∑*_i_β_i_* ·*r_ij_*, where *β_i_* is the estimated effect size of the *i^th^* CpG obtained from the regularized regression, *r_ij_* is the residuals of CpG, for individual *j*. The scores were standardized to have a mean of 0 and a standard deviation (SD) of 1, referred to as CpG-standardized scores. Separate CpG-standardized scores were obtained for the CpGs associated with MHC and MSS.

#### Association analyses with all-cause mortality and CVD

The Cox proportional hazard model was fitted to evaluate the associations of all-cause mortality and CVD with the CpG-standardized scores for both MHC and MSS. The base model includes age and sex as covariates. The second model additionally accounted for smoking status (never, former, and current). Smoking status was used instead of smoking score because the former has traditionally been included in association analyses of all-cause mortality or CVD as the outcome. Additionally, the smoking score variable was not available in the replication cohort. The multi-covariate model included age, sex, smoking status, BMI, systolic blood pressure (SBP), use of antihypertensive medication, total cholesterol, high-density cholesterol, diabetes, and use of lipid-lowering medication. We conducted race- and cohort-specific association analyses in JHS, MESA, and WHI. Since FHS was used as the training cohort to select CpGs, a meta-analysis was performed using the random effects inverse variance method, excluding FHS for internal validation.

#### External replication

The Health and Retirement Study (HRS), established in 1992, recruits participants aged 50 and older, along with their spouses, to investigate factors related to aging.^68^ HRS was utilized as an independent replication cohort to evaluate the association between the CpG-standardized scores and all-cause mortality as well as CVD. In HRS, DNAm was measured using Illumina HumanMethylationEPIC BeadChip (**Supplemental Material**). Race/ethnicity-specific association analyses were conducted in self-reported White (n=2506), Black (n=612), and Hispanic participants (n=532). The proportional hazards assumption was examined and found to be met in all analyses.

URLs

MRC-IEU EWAS Catalog: http://www.ewascatalog.org

DAVID: https://david.ncifcrf.gov/

## Supporting information

Supplemental Methods and Results

## Data Availability

All data produced are available online at https://www.ncbi.nlm.nih.gov/gap/

## TOPMed Acknowledgement

The Atherosclerosis Risk in Communities study (ARIC) has been funded in whole or in part with federal funds from the National Heart, Lung, and Blood Institute, National Institute of Health, Department of Health and Human Services, under contract numbers (HHSN268201700001I, HHSN268201700002I, HHSN268201700003I, HHSN268201700004I, and HHSN268201700005I). The authors thank the staff and participants of the ARIC study for their important contributions. WGS for “NHLBI TOPMed: Atherosclerosis Risk in Communities” (phs001211.v3.p2.c1) was performed at the Baylor College of Medicine Human Genome Sequencing Center (3U54HG003273-12S2 / HHSN268201500015C). Core support including centralized genomic read mapping and genotype calling, along with variant quality metrics and filtering were provided by the TOPMed Informatics Research Center (3R01HL-117626-02S1; contract HHSN268201800002I). Core support including phenotype harmonization, data management, sample-identity QC, and general program coordination were provided by the TOPMed Data Coordinating Center (R01HL-120393; U01HL-120393; contract HHSN268201800001I). We gratefully acknowledge the studies and participants who provided biological samples and data for TOPMed.

The Coronary Artery Risk Development in Young Adults Study (CARDIA) for the NHLBI TOPMed program: CARDIA (phs001612) was performed at the Baylor Human Genome Sequencing Center (HHSN268201600033I). Centralized read mapping and genotype calling, along with variant quality metrics and filtering were provided by the TOPMed Informatics Research Center (3R01HL-117626-02S1). Phenotype harmonization, data management, sample-identity QC, and general study coordination, were provided by the TOPMed Data Coordinating Center (3R01HL-120393-02S1). We gratefully acknowledge the studies and participants who provided biological samples and data for TOPMed. The Coronary Artery Risk Development in Young Adults Study (CARDIA) is conducted and supported by the National Heart, Lung, and Blood Institute (NHLBI) in collaboration with the University of Alabama at Birmingham (75N92023D00002 & 75N92023D00005), Northwestern University (75N92023D00004), University of Minnesota (75N92023D00006), and Kaiser Foundation Research Institute (75N92023D00003). The authors also wish to thank the staffs and participants of the CARDIA.

The Framingham Heart Study (FHS) is conducted and supported by the National Heart, Lung, and Blood Institute (NHLBI) in collaboration with Boston University (Contract No. N01-HC-25195, HHSN268201500001I and 75N92019D00031). This manuscript was not prepared in collaboration with investigators of the Framingham Heart Study and does not necessarily reflect the opinions or views of the Framingham Heart Study, Boston University, or NHLBI.

The Genetic Epidemiology Network of Arteriopathy (GENOA) is supported by the National Heart, Lung and Blood Institute (U01HL054457, U01HL054464, U01HL054481, R01HL119443, and R01HL087660) of the National Institutes of Health. Sequencing for the GENOA (phs001345.v1.p1) was performed by the University of Washington Northwest Genomics Center (3R01HL055673-18S1) and at the Broad Institute of MIT and Harvard (HHSN268201500014C)). The authors also wish to thank the staff and participants of GENOA.

The Jackson Heart Study (JHS) is supported and conducted in collaboration with Jackson State University (HHSN268201800013I), Tougaloo College (HHSN268201800014I), the Mississippi State Department of Health (HHSN268201800015I/HHSN26800001) and the University of Mississippi Medical Center (HHSN268201800010I, HHSN268201800011I and HHSN268201800012I) contracts from the National Heart, Lung, and Blood Institute (NHLBI) and the National Institute for Minority Health and Health Disparities (NIMHD). The authors also wish to thank the staffs and participants of the JHS.

The Multi-Ethnic Study of Atherosclerosis Study (MESA) Cohort : WGS for the TOPMed program was supported by the NHLBI. WGS for the NHLBI’s TOPMed (phs001416.v1.p1) was performed at the Broad Institute of MIT and Harvard (3U54HG003067-13S1). Centralized read mapping and genotype calling, along with variant quality metrics and filtering were provided by the TOPMed Informatics Research Center (3R01HL-117626-02S1). Phenotype harmonization, data management, sample-identity QC, and general study coordination, were provided by the TOPMed Data Coordinating Center (3R01HL-120393-02S1). MESA and the MESA SHARe project (phs001416.v1.p1) are conducted and supported by the NHLBI in collaboration with MESA investigators. Support for MESA is provided by contracts 75N92020D00001, HHSN268201500003I, N01-HC-95159, 75N92020D00005, N01-HC-95160, 75N92020D00002, N01-HC-95161, 75N92020D00003, N01-HC-95162, 75N92020D00006, N01-HC-95163, 75N92020D00004, N01-HC-95164, 75N92020D00007, N01-HC-95165, N01-HC-95166, N01-HC-95167, N01-HC-95168, N01-HC-95169, UL1-TR-000040, UL1-TR-001079, UL1-TR-001420, UL1-TR-001881, and DK063491. Funding for SHARe genotyping was provided by NHLBI Contract N02-HL-64278. Genotyping was performed at Affymetrix (Santa Clara, California, USA) and the Broad Institute of Harvard and MIT (Boston, Massachusetts, USA) using the Affymetrix Genome-Wide Human SNP Array 6.0. Also supported in part by the National Center for Advancing Translational Sciences, CTSI grant UL1TR001881, and the National Institute of Diabetes and Digestive and Kidney Disease Diabetes Research Center (DRC) grant DK063491 to the Southern California Diabetes Endocrinology Research Center.

The Women’s Health Initiative (WHI) program is funded by the National Heart, Lung, and Blood Institute, National Institutes of Health, U.S. Department of Health and Human Services through contracts HHSN268201600018C, HHSN268201600001C, HHSN268201600002C, HHSN268201600003C, and HHSN268201600004C. The TOPMed WHI (phs001237) were supported by HHSN268201500014C and HHSN268201600034I (Broad Institute, WGS, RNAseq and metabolomics), HHSN268201600038I (Keck MGC, Methylomics). Scientific Computing Infrastructure at Fred Hutch was funded by ORIP grant S10OD02868. We thank all WHI participants for their dedications and contributions.

