## Supplemental Methods and Results for "Epigenome-wide Association Analysis of Mitochondrial Heteroplasmy Provides Insight into Molecular Mechanisms of Disease"

Lai. et al.

**SUPPLEMENTARY METHODS**

**Study participants**

This study consists of participants from seven cohorts: ARIC, CARDIA, FHS, GENOA, JHS, MESA, WHI. The Institutional Review Board (IRB) of each institute approved the study, and informed consent were received from all participants.

**Disclaimer -** The views expressed in this manuscript are those of the authors and do not necessarily represent the views of the National Heart, Lung, and Blood Institute; the National Institutes of Health; or the U.S. Department of Health and Human Services.

*Atherosclerosis Risk in Communities study (ARIC)*:

ARIC is a prospective heart health study that investigates clinical outcomes in participants from four U.S. communities: Forsyth County, NC; Jackson, MS; the northwest suburbs of Minneapolis, MN; and Washington County, MD. The event adjudication (to the end of 2017) focused on cardiovascular disease outcomes, and conducted a committee review of death certificates, hospital records and phone interview reports. Gentra Puregene Blood Kit (Qiagen) was used for the purification of DNA from buffy coat in whole blood. This study includes the whole genome sequencing results from 3074 participants.

Cohort acknowledgement: The National Heart, Lung, and Blood Institute (NHLBI) sponsored the whole genome sequencing (WGS) for the Trans-Omics in Precision Medicine (TOPMed). WGS for ARIC (phs001211) was performed by the Baylor College of Medicine Human Genome Sequencing Center, supported by HHSN268201500015C and 3U54HG003273-12S2, the Broad Institute for MIT and Harvard (3R01HL092577- 06S1), and the National Human Genome Research Institute grants U54 HG003273 and UM1 HG008898. TOPMed Informatics Research Center (3R01HL-117626-02S1) provided centralized read mapping, genotype calling, variant quality metrics, and filtering. TOPMed Data Coordinating Center (3R01HL- 120393- 02S1) carried out study coordination, data management, phenotype harmonization, and sample-identity QC. The National Eye Institute (NEI), National Human Genome Research Institute (NHGRI) and the NHLBI sponsored the Genome Sequencing Program (GSP). The GSP Coordinating Center (U24 HG008956) provided study coordination and cross program scientific initiatives. Support for the Centers for Common Disease Genomics (CCDG) program was provided by the NHGRI and NHLBI. This research was sponsored by the NIH grant R01HL131573 awarded to Dr. Dan Arking. The ARIC study has also been funded by the National Heart, Lung, and Blood Institute, National Institutes of Health, Department of Health, and Human through contracts HHSN268201700001I, HHSN268201700002I, HHSN268201700003I, HHSN268201700004I and HHSN268201700005I. Funds were also provided by 5RC2HL102419 and R01NS087541 for obtaining DNA methylation data. “Building on GWAS for NHLBI-diseases: the U.S. CHARGE consortium” was sponsored by NIH and the American Recovery and Reinvestment Act of 2009 (ARRA) (5RC2HL102419). The authors gratefully acknowledge all staff and study participants of the ARIC study.

*Coronary Artery Risk Development in Young Adults (CARDIA)*:

The Coronary Artery Risk Development in Young Adults (CARDIA) is a prospective cohort study Initiated in 1985-86. The CARDIA was aimed to investigate lifestyle and other factors that influence CVD during young adulthood. The study participants aged 18-30 years were recruited and examined in four urban areas: Birmingham, Alabama; Chicago, Illinois; Minneapolis, Minnesota, and Oakland, California. The initial examination included 5,115 Black and White women and men. The carefully standardized measurements were conducted on study participants to investigate major risk factors, such as blood pressure, cholesterol and other lipids, and glucose, for CVD. Physical measurements such as weight and body composition as well as lifestyle factors were also collected. This study included 2257 Black and White participants who had both DNA methylation and WGS.

Cohort acknowledgement: The Coronary Artery Risk Development in Young Adults Study (CARDIA) is conducted and supported by the National Heart, Lung, and Blood Institute (NHLBI) in collaboration with the University of Alabama at Birmingham (75N92023D00002 & 75N92023D00005), Northwestern University (75N92023D00004), University of Minnesota (75N92023D00006), and Kaiser Foundation Research Institute (75N92023D00003).CARDIA was also supported in part by the Intramural Research Program of the National Institute on Aging (NIA) and an intra-agency agreement between NIA and NHLBI (AG0005). The DNA methylation laboratory work and analytic component were funded by the American Heart Association (17SFRN33700278 and 14SFRN20790000, Northwestern University, to Dr Hou) and NIA R21AG068955 (to Drs Liu and Zheng).

*The Framingham Heart Study (FHS)*:

The Framingham Heart Study is a single-site, generational study that investigates the risk factors contributing to cardiovascular disease. TOPMed administrated the whole genome sequencing of 4,196 FHS participants. Among these samples, there were 376 first generation participants, 2218 participants in the Offspring cohort, 95 participants in the New Offspring Spouse cohort, and 1507 participants from the Third Generation. Exam on first generation participants were performed every two years. The Offspring cohort, recruited in 1971, were examined every four to eight years. The Offspring cohort included the children of the first generation and the spouses of the children. The Third Generation participants and an additional 100 spouses of the Offspring were recruited during 2002-2005. Three examinations were performed on the Third Generation participants and the spouses of the second generation participants. Informed consents were provided by all participants. DNA methylation levels of the 1788 FHS participants were measured at the same time of WGS.

Cohort acknowledgement: Whole genome sequencing for Framingham Heart Study (phs000974) was performed by Broad Institute of MIT and Harvard (3R01HL092577-06S1 and 3U54HG003067-12S2). This research is supported by the National Heart, Lung and Blood Institute and grant supplement R01-HL092577-06S1 through contracts NO1-HC-25195, HHSN268201500001I and 75N92019D00031. We thank the FHS study participants for their dedication. R01AG059727. P.W. supported X.S. and C.L., and NHLBI intramural funds supported T.H., R.J., and D.L. The DNA methylation data was provided by NHLBI’s Systems Approach to Biomarker Research in Cardiovascular Disease Initiative (SABRe CVD Initiative) and sponsored by the NIH intramural fund.

*The Genetic Epidemiology Network of Arteriopathy (GENOA)*:

The GENOA study investigates hypertensive sibships. At least two siblings of the participants were diagnosed of essential hypertension prior to the age of 60. The initial study phase (1995-2000) recruited 1854 African Americans from Jackson, Mississippi and 1583 non-Hispanic white adults from Rochester, Minnesota. Besides hypertensive siblings, normotensive members within the sibship were also included. The second phase of the GENOA study recruited additional participants recruited 2000-2005. The biological samples in GENOA, as well as information such as generic and anthropometric data, contributes to the understanding the development of heart, brain, peripheral arteries, and kidney disease. A total of 797 African American participants with both the WGS data and Phase I DNA methylation data are included in our study. Informed consents were provided by all participants.

Cohort acknowledgement: This study is supported by the National Heart, Lung, and Blood Institute of the National Institutes of Health (U01HL054457, U01HL054464, U01HL054481, R01HL087660, RC1HL100185, R01HL119443, and R01HL133221). Whole genome sequencing for GENOA (phs001345.v1.p1) was performed at the University of Washington Northwest Genomics Center. National Heart, Lung, and Blood Institute. This research is also supported by the National Heart, Lung and Blood Institute and grant supplement R01-3R01HL055673-18S1 and the Broad Institute of MIT and Harvard (HHSN268201500014C). The authors thank all the staff and participants of the GENOA study.

*The Jackson Heart Study (JHS)*:

The JHS is one of the largest prospective, epidemiologic investigation of CVD among African Americans. The study participants were recruited in the three counties (Hinds, Madison, and Rankin) in the Jackson, Mississippi metropolitan area. Regular health were conducted to collect data and biologic materials from 5,306 participants. Among these participants included a nested family cohort of 1,498 members of 264 families. The initial age was 35-84 years for the unrelated cohort; participants >21 years old were recruited in the family cohort. Extensive medical, social history, and an array of physical and biochemical measurements and diagnostic procedures were measured during a baseline examination (2000-2004), two follow-up examinations (2005-2008 and 2009-2012), and ancillary studies. Genomic DNA samples were collected during the first two examinations. A total of 3482 participants provided consent for genetic studies and broad sharing of genetic data. Whole genome sequence data are available for 3,406 participants after quality control. To identify intervening clinical events, follow-up information on vital status, major illnesses or injuries, and hospitalizations were conducted annually by phone. Medical records of cardiovascular disease related hospitalizations and death certificates are abstracted and used for adjudication of cardiovascular events and related deaths.

Cohort acknowledgement/support: The Jackson Heart Study (JHS) is supported and conducted in collaboration with Jackson State University (HHSN268201800013I), Tougaloo College (HHSN268201800014I), the Mississippi State Department of Health (HHSN268201800015I) and the University of Mississippi Medical Center (HHSN268201800010I, HHSN268201800011I and HHSN268201800012I) contracts from the National Heart, Lung, and Blood Institute (NHLBI) and the National Institute on Minority Health and Health Disparities (NIMHD). The authors also wish to thank the staffs and participants of the JHS. Molecular data for the Trans-Omics in Precision Medicine (TOPMed) program was supported by the National Heart, Lung, and Blood Institute (NHLBI). Genome sequencing for “NHLBI TOPMed: The Jackson Heart Study” (phs000964.v1.p1) was performed at the Northwest Genomics Center (HHSN268201100037C). Core support including centralized genomic read mapping and genotype calling, along with variant quality metrics and filtering were provided by the TOPMed Informatics Research Center (3R01HL-117626-02S1; contract HHSN268201800002I). Core support including phenotype harmonization, data management, sample-identity QC, and general program coordination were provided by the TOPMed Data Coordinating Center (R01HL-120393; U01HL-120393; contract HHSN268201800001I). Laura Raffield was also supported by the National Center for Advancing Translational Sciences, National Institutes of Health, through Grant KL2TR002490 (LMR). We gratefully acknowledge the studies and participants who provided biological samples and data for TOPMed.

*Multi-Ethnic Study of Atherosclerosis Study (MESA)*:

MESA is a population-based study of subclinical cardiovascular disease and clinically overt cardiovascular disease. The MESA study includes 6814 participants aged 45 to 84 without known clinical cardiovascular disease at baseline, recruited from six field centers. Informed consents were provided by all participants. Gentra Puregene Blood Kit was used to extract DNA from peripheral leukocytes (obtained in exam 1). Event adjudication (2000-2015) involved a committee review of death certificates, hospital records and phone interview reports. Our study included 854 MESA participants with WGS and DNA methylation data.

Cohort acknowledgement: Whole genome sequencing (WGS) for the Trans-Omics in Precision Medicine (TOPMed) program was supported by the National Heart, Lung, and Blood Institute (NHLBI). WGS for “NHLBI TOPMed: Multi-Ethnic Study of Atherosclerosis (MESA)” (phs001416.v1.p1) was performed at the Broad Institute of MIT and Harvard (3U54HG003067-13S1). Centralized read mapping and genotype calling, along with variant quality metrics and filtering were provided by the TOPMed Informatics Research Center (3R01HL-117626-02S1). Phenotype harmonization, data management, sample-identity QC, and general study coordination, were provided by the TOPMed Data Coordinating Center (3R01HL-120393-02S1), and TOPMed MESA Multi-Omics (HHSN2682015000031/HSN26800004). The MESA projects are conducted and supported by the National Heart, Lung, and Blood Institute (NHLBI) in collaboration with MESA investigators. Support for the Multi-Ethnic Study of Atherosclerosis (MESA) projects are conducted and supported by the National Heart, Lung, and Blood Institute (NHLBI) in collaboration with MESA investigators. Support for MESA is provided by contracts 75N92020D00001, HHSN268201500003I, N01-HC-95159, 75N92020D00005, N01-HC-95160, 75N92020D00002, N01-HC-95161, 75N92020D00003, N01-HC-95162, 75N92020D00006, N01-HC-95163, 75N92020D00004, N01-HC-95164, 75N92020D00007, N01-HC-95165, N01-HC-95166, N01-HC-95167, N01-HC-95168, N01-HC-95169, UL1-TR-000040, UL1-TR-001079, UL1-TR-001420, UL1TR001881, DK063491, and R01HL105756. The authors thank the other investigators, the staff, and the participants of the MESA study for their valuable contributions. A fill list of participating MESA investigators and institutes can be found at http://www.mesa-nhlbi.org. This study was also supported in part by the NIH contracts R01AA082263 and R01HL155569.

*The Women’s Health Initiative (WHI)*:

WHI is a prospective health study designed to examine chronic diseases that are responsible for major mortality and disability in postmenopausal women and develop prevention strategies. In the initial stage, 161,808 women of age 50-79 were recruited (1993-1997) to construct a sample consisting of socio-demographically diverse population with subjects from racial or ethnic minority groups in a way that is consistent with the proportion of total minority women population in the U.S. within that age range. The WHI study included both the randomized Clinical Trials (CT; N=68,132) and the Observational Study (OS; N = 93,676). The clinical trials had three components that overlapped with each other: Hormone Therapy Trials, Dietary Modification Trial, and Calcium and Vitamin D Trial. Each of these three trials was aimed to compare the administered therapies between the controlled and the randomized experiment. The parallel observational study investigated biomarkers and risk factors in association with chronic diseases. The Hormone Therapy Trials ended around 2005. However, the WHI-CT and WHI-OS cohorts continued to perform follow-up exams for more than 25 years, in which various types of data such as clinical outcomes and risk factor measurements were collected. Our study included 521 WHI participants with WGS and DNA methylation data.

Cohort acknowledgement: The WHI study was sponsored by U.S. Department of Health and Human Services, and the National Heart, Lung, and Blood Institute. Support was provided by contracts 75N92021D00001, 75N92021D00002, 75N92021D00003, 75N92021D00004, 75N92021D00005. We thank all WHI participants for their dedications and contributions.. The ORIP grant S10OD02868 funded the Scientific Computing Infrastructure at Fred Hutch. WGS for WHI (phs001237) was supported by Broad Institute through contracts HHSN268201500014C, HHSN268201600034I (WGS, RNAseq and metabolomics), and HHSN268201600038I (Keck MGC, Methylomics). We also thank all participants for their contribution to the WHI study.

*Health and Retirement Study (HRS):*

The Health and Retirement Study (HRS) is a longitudinal survey of a representative sample of Americans over the age of 50 (Sonnega, 2014). Over 42,000 persons in 26,000 households have been interviewed since 1992. The study interviews respondents every two years about income and wealth, health and use of health services, work and retirement, and family connections. The sample is refreshed periodically with a new cohort of respondents to offset attrition and death and maintain ~20,000 individuals per sample wave. Starting in 2006, half of the core sample is randomly assigned to a face-to-face interview enhanced with physical and biological measures and a mail-back psychosocial questionnaire. The other half is interviewed by telephone. Each interview mode then alternates between the two half-samples in subsequent waves.

Cohort acknowledgement: HRS is supported by the National Institute on Aging (NIA U01AG009740). The HRS DNA methylation was measured by the University of Minnesota Genomics Center (UMGC). Sample coordination, plate design, and quality control was performed by the University of Minnesota Advanced Research and Diagnostics Laboratory (ARDL).

**DNA methylation profiling, processing, and quality control**

*ARIC*: Our study included the WGS and DNA methylation data from 3074 ARIC participants who self-reported as White or Black participants. Race-specific data normalization was done using BMIQ and then processed by Noob. We excluded individuals with low DNA sample pass rate, possible gender, or genotype mismatch. CpG sites with low detection rate were also excluded from analyses ($\frac{number of probes with detection p-value<0.01}{total number of probes in array}$ < 95%).

*CARDIA*: DNA methylation was analyzed using the Infinium MethylationEPIC BeadChip (EPIC array) in whole blood.^1^ Quality control and preprocessing were performed with the R package ENmix using default settings. Low-quality methylation measurements were identified and excluded based on detection p-values <1.0E-6 or bead counts <3. We removed 6,209 CpG sites with a detection rate <95% and 87 samples with >5% low-quality measurements or very low bisulfite conversion probe intensity. After this, 95 extreme outlier samples, defined by Tukey’s method, were also excluded. The remaining samples underwent ENmix preprocessing, including background correction and dye bias correction using RELIC. M and U intensities were separately quantile-normalized for Infinium I and II probes. Low-quality methylation values and extreme β-value outliers were set as missing. The final dataset included 841,639 autosomal CpG probes and 1,999 samples from 1,118 participants (1,042 from Y15, 957 from Y20, and 881 from both examinations).

*FHS*: Our study included 1788 participants whose WGS and DNA methylation data was measured at the same exams. DNA methylation measurement on the exam 8 Offspring subjects (N=2,846; 2005-2008) and exam 2 Generation-3 individuals (N=1,549; 2008-2011) was sponsored by the NHLBI intramural funds awarded to Daniel Levy. DNA extraction from peripheral whole blood followed the standard procedures. Gentra Puregene DNA extraction kit (Qiagen, Venlo, Netherlands) was used for genomic DNA extraction from buffy coat. EZ DNA Methylation Kit (Zymo Research, Irvine, CA) was used for bisulfite conversion of DNA. DNA methylation levels were measured using Infinium HumanMethylation450 BeadChip (Illumina Inc, San Diego, CA). Three laboratories contributed to this analysis. Methylation levels of 576 samples, which were included in a previous case-control study of cardiovascular disease, was measured at one lab. The remaining 2,270 samples from the Offspring cohort were analyzed at another lab. Illumina (Illumina Inc, San Diego, CA) performed measurements on the rest of the samples. Total probe intensity and the methylated probe intensity were obtained using Illumina Genome Studio (version 2011.1) and methylation module (version 1.9.0). The methylation beta was defined as follows: $= \frac{M}{M+U+100}$ . Here M was the methylated signal and U was the unmethylated signal. To restrict technical artifacts, we used the DASEN method in wateRmelon/Rin software to normalize the resulting betas. In conducting QC, multidimensional scaling analysis (MDS) was performed to identify outliers and gender mismatch. To ensure consistency, 65 overlapping SNPs were compared to previous genotyping or 1000 Genome imputation and the Illumina 450K methylation array. We removed low quality probes with high missing rate (>20%), mapping to different locations, involving SNPs at CpG sites (MAF>5% in EUR 1000G), or having at most 10 base pair Single Base Extension. Samples were excluded if they had high missing rate (>1%), identified as MDS outliers, or was a poor match to the SNP genotype.

*GENOA*: AutoGen FlexStar (AutoGen, Holliston, MA) was used to extract genomic DNA from peripheral blood leukocytes collected during Phase I (N=1106) and Phase 2 (N=304) of the GENOA study. The EZ DNA Methylation Kit (Zymo Research, Irvine, CA) was used for bisulfite conversion of the obtained DNA. Methylation level was measured using the Illumina Infinium HumanMethylationEPIC BeadChip (Illumina Inc, San Diego, CA). Density plot, using shinyMethyl R package, was generated to find gender mismatch and outliers in the raw intensity data. Function QCinfo() in the ENmix R package was used to identify incomplete bisulfite conversions, and related samples were removed. Samples also went through misalignment check using the 59 SNP probes implemented in the EPIC chip. Background correction was performed using Minfi R package, and data normalization was done using Noob. To adjust for probe-type bias, we applied the regression on correlated probes (RCP) method. We excluded the probes and samples with detection rate less than 0.01. A total of 857,121 remaining probes were from 1100 Phase I samples and Phase II 294 samples. The Houseman method was then used to assess the white blood cell counts.

*JHS*: The Jackson Heart Study methylation level was measured by the Illumina EPIC (850k) array. The data was normalized using the minfi R package, and samples that did not pass quality control were removed by Steve Horvath's group prior to adjustment for technical variation/batch effects. Noob normalization and generation of methylation beta values from both ASN0104 and ASN0148 was performed by Steve Horvath’s lab. Sample outlier were identified based on hierarchical clustering and removed. The dataset used in this study ‘jhs_betas_combat_adjusted' was adjusted for known batch effects (group, plate, well) using the SVA R package v3.30.1 ComBAT function. This method uses surrogate variable analysis to perform a global data adjustment based on batch effects. In this dataset, there are 866836 probes and 1706 individuals. JHS participants have approximately 83% mean similarity to 1000G AFR reference panels based on prior work, but no participants were excluded based on ancestry proportions.

*MESA*: Our study included WGS and DNA methylation data from 174 participants who self-identified as Black or African American, 286 as Hispanic/Latino, and 394 as White. NHLBI’s TOPMed program (phs001416.v1.p1), contract HHSN268201600034I, sponsored the DNA methylation measurements. Methylation level was assessed using the Illumina Infinium HumanMethylationEPIC BeadChip. The methylation beta was estimated as follows: $= \frac{M}{M+U+100}$ . Here M was the methylated signal and U was the unmethylated signal, Watermelon package and minfi(v1.22.1) package were used to process the raw data (.idat). To perform background correction and normalization, we used normal-exponential convolution with out-of-band Infinium I probes (noob). The sample concordance were checked by comparing the genotypes identified in previous genotyping arrays to the 59 SNPs in the methylation data. (Details of the profiling and quality control procedure can be found in https://www.nhlbiwgs.org/sites/default/files/TOPMed_Methylation_array_pipeline_COREyr3.pdf) Sex mismatch samples were removed, and probes with detection p-values > 0.05 were assigned value of NA.

*WHI*: Our study included participants whose WGS and DNA methylation data was measured at the same exams. Gentra Puregene DNA extraction kit (Qiagen, Venlo, Netherlands) was used for genomic DNA extraction from peripheral whole blood. EZ DNA Methylation Kit (Zymo Research, Irvine, CA) was used to perform DNA bisulfite conversion. DNA methylation level was assessed using the Infinium HumanMethylation450 BeadChip (Illumina Inc, San Diego, CA). We excluded probes with detection p-values greater than 0.01 in more than one percent of all samples, CpG sites on Y chromosome, less than 3 bead count in more than ten percent of the samples, and also the ones that had measurements on non-CpG sites. The beta-mixture quantile normalization in BMIQ was used to perform normalization. The resulting methylation data is the adjusted ratio of the methylated allele to the sum of the methylated and unmethylated allele.

*Health and Retirement Study (HRS):* As part of the Venous Blood Study (VBS), an ancillary study of HRS, venous blood was collected in the 2016 wave. For a total of 4,103 respondents, DNA methylation was assessed from the blood samples using the Illumina HumanMethylationEPIC BeadChip. Data were imported using the minfi R package (Aryee, 2014). Sex mismatches and those with an average median intensity <8.5 were removed. A detection p-value<0.01 was used to remove samples and probes with detection rate <5%. Sample identity was assessed using 59 SNP probes on the EPIC chip, but no additional samples were removed. After exclusions, a total of 836,660 probes in 4,018 samples were available for analysis.

**DNA methylation-based smoking score**

Smoking is strongly associated with numerous DNA methylation sites, leading to distinct methylation profiles between individuals who currently smoke and those who never smoke. ^2^ Smoking is also strongly associated with mtDNA heteroplasmy^3^. The EpiSmokEr R package has been developed to estimate smoking status based on 183 CpGs^4^. We calculated the methylation-based smoking scores for all participants using the function epismoker() with the method “SSc” in the epiSmokEr package. Compared to self-reported smoking status, smoking scores may provide a more objective assessment for a person’s smoking status and better reflect smoking history (e.g., recall bias, second-hand smoking or missing data due to reluctance of reporting).^4^ The calculated smoking scores were used in the primary analysis of DNAm and mtDNA heteroplasmy.

**Comparison of associations of heteroplasmy and DNA methylation between Black and White individuals**

To explore race-specific association between CpGs and heteroplasmy, we conducted meta-analyses in American Whites only (n=5104) (**Supplemental Figure 3**) and African Americans only samples (n=5882) (**Supplemental Figure 4**). With a relaxed threshold(*p* < 1e-4), we observed much fewer significant CpGs in African Americans (n=44 CpGs with MSS and 21 with MHC) than American Whites (n=1295 CpGs with MSS and 524 with MHC). We compared the top CpGs identified between racial groups. Using the CpGs identified in African Americans (*p* < 1e-4) as the reference set, we observed moderate (Pearson correlation r = 0.54 of the 44 CpGs with MSS) to high (Pearson correlation r = 0.91 of the 21 CpGs with MHC) correlations in the beta estimates of the CpGs (**Supplemental Figure 5, 6**). Conversely, when using the CpGs identified in American Whites as the reference set, we found low correlations (Pearson correlation r= 0.22 for the 1295 CpGs and r=0.30 for the 524 CpGs) in the estimates of the CpGs (**Supplemental Figure 5, 6**).

**Supplemental Tables and Figures.**

**Supplemental Table 1**. Cohort characteristics: Self-reported American White and Hispanic American participants

| **Variable** | **ARIC EA**  **(n=960)** | **CARDIA EA (N= 1268)** | **FHS EA (N= 1788 )** | **MESA EA (N= 394)** | **MESA HIS (N= 286)** | **WHI BAA23 EA (N= 70)** | **WHI EMPC EA (N= 338)** |
| --- | --- | --- | --- | --- | --- | --- | --- |
| Age at blood draw,  yrs, mean (±SD) | 62.08 (5.1) | 47.6 (4.9) | 62.8 (12.6) | 60.9 (9.8) | 58.7 (9.4) | 67.1 (6.1) | 65.80 (6.8) |
| Women, n(%) | 563 (58.6%) | 688 (54.3%) | 957 (53.5%) | 197 (50%) | 159 (55.6%) | 70 (100%) | 338 (100%) |
| Smoking score, median  (Q1, Q3) | - 3.2 ( - 5.4, 0.8) | -8.0 (-10.0, -5.0) | 0.3 (-2.0,3.3) | -5.1 (-7.1,-2.8) | -5.7 (-7.4,-3.7) | -3.7 (-6.1, -0.7) | -1.8 (-4.2,1.7) |
| Smoking status, n (%) |  |  |  |  |  |  |  |
| Never smoker | 410 (42.7%) | 771 (60.8%) | 1118 (62.5%) | 162 (41.1%) | 150 (52.5%) |  |  |
| Former smoker | 382 (39.8%) | 325 (25.6%) | 516 (28.7%) | 179 (45.4%) | 101 (35.3%) | 59 (84.3%)* | 306 (90.5%) |
| Current smoker | 168 (17.5%) | 172 (13.6%) | 154 (8.6%) | 53 (13.5%) | 35 (12.2%) | 11 (15.7%) | 32 ( 9.5%) |
| Heteroplasmy |  |  |  |  |  |  |  |
| Prevalence  (>1 MH count), n (%) | 319 (33.2%) | 357 (28.2%) | 587 (32.8%) | 119 (30.2%) | 99 (34.6%) | 17 (24.3%) | 102 (30.2%) |
| 0.01-0.25 MSS, n (%) | 125 (13.0%) | 151 (11.9%) | 148 (8.3%) | 49 (12.4%) | 37 (12.9%) | 6 (8.6%) | 36 (10.7%) |
| 0.251-0.5 MSS, n (%) | 44 (4.6%) | 45 (3.5%) | 103 (5.8%) | 13 (3.3%) | 12 (4.2%) | 2 (2.9%) | 9 (2.7%) |
| >0.5 MSS, n(%) | 79 (8.2%) | 68 (5.4%) | 190 (10.6%) | 26 (6.6%) | 23 (8.0%) | 4 (5.7%) | 25 (7.4%) |

ARIC, Atherosclerosis Risk in Communities. CARDIA, Coronary Artery Risk Development in Young Adults. FHS, Framingham Heart Study. GENOA, Genetic Epidemiology Network of Arteriopathy. MESA, Multi-Ethnic Study of Atherosclerosis. WHI, Women’s Health Initiative.

**Supplemental Table 2**. Cohort characteristics: Self-reported American Black participants

| **Variable** | **ARIC AA**  **(n=2114)** | **CARDIA AA (N= 989)** | **GENOA AA (N= 786)** | **JHS AA (N= 1706)** | **MESA AA (N= 174)** | **WHI EMPC AA (N= 113)** |
| --- | --- | --- | --- | --- | --- | --- |
| Age at blood draw,  yrs, mean (±SD) | 56.6 (5.9) | 46.5 (5.4) | 61.1 (10.4) | 56.18 (12.33) | 60.7 (9.6) | 65.34 (6.79) |
| Women, n (%) | 1327 (62.8%) | 626 (63.3%) | 565 (71.9%) | 1162 (68.11%) | 101 (58.05%) | 113 (100%) |
| Smoking score, median  (Q1, Q3) | - 2.1 (- 5.1, 3.0) | -6.0 (-9.0, -2.0) | -8.6 (-10.7,-5.8) | -4.5 (-6.4,-1.8) | -4.6 (-6.6,-2.3) | -1.3 (-3.3,1.6) |
| Smoking status, n(%) | | | | | | |
| Never smoker | 919 (43.5%) | 618 (62.5%) | 485 (61.7%) | 1108 (65.0%) | 80 (46.0%) |  |
| Former smoker | 630 (29.8%) | 147 (14.9%) | 171 (21.8%) | 355 (20.8%) | 60 (34.5%) | 102 (90.3%) |
| Current smoker | 565 (26.7%) | 224 (22.6%) | 130 (16.5%) | 243 (14.3%) | 34 (19.5%) | 11 (9.7%) |
| Heteroplasmy |  |  |  |  |  |  |
| Prevalence  (>1 MH count), n(%) | 678 (32.1%) | 328 (33.2%) | 318 (40.5%) | 544 (31.9%) | 63 (36.2%) | 29 (25.7%) |
| 0.01-0.25 MSS, n (%) | 276 (13.1%) | 145 (14.7%) | 146 (18.8%) | 198 (11.6%) | 32 (18.4%) | 9 (8.0%) |
| 0.251-0.5 MSS, n (%) | 92 (4.4%) | 39 (3.9%) | 40 (5.1%) | 69 (4.0%) | 9 (5.2%) | 5 (4.4%) |
| >0.5 MSS, n(%) | 155 (7.3%) | 73 (7.4%) | 85 (108%) | 120 (7.0%) | 11 (6.3%) | 6 (5.3%) |

ARIC, Atherosclerosis Risk in Communities. CARDIA, Coronary Artery Risk Development in Young Adults. GENOA, Genetic Epidemiology Network of Arteriopathy. JHS, Jackson Heart Study. MESA, Multi-Ethnic Study of Atherosclerosis. WHI, Women’s Health Initiative

**Supplemental Table 3**. Platform and sequencing information by cohort and race.

| **Study** | **n** | | **DNA Methylation**  **Platform^1^** | **Sequencing Platform** |
| --- | --- | --- | --- | --- |
| **African American (n = 5882)** | | | | |
| ARIC | 2114 | | 450K BeadChip | WGS |
| CARDIA | 989 | | MethylationEPIC BeadChip | WGS |
| GENOA | 786 | | MethylationEPIC BeadChip | WGS |
| JHS | 1706 | | MethylationEPIC BeadChip | WGS |
| MESA | 174 | | MethylationEPIC BeadChip | WGS |
| WHI EMPC | 113 | | HumanMethylation 450K BeadChip | WGS |
| **European American (n = 4818)** | | | | |
| ARIC | 960 | | 450K BeadChip | WGS |
| CARDIA | 1268 | | EPIC BeadChip | WGS |
| FHS | 1788 | | 450K BeadChip | WGS |
| MESA | 394 | | EPIC BeadChip | WGS |
| WHI BAA23 | | 70 | 450K BeadChip | WGS |
| WHI EMPC | 338 | | 450K BeadChip | WGS |
| **Hispanic American (n=286)** | | | | |
| MESA | 286 | | MethylationEPIC BeadChip | WGS |

^1^450K BeadChip, Infinium HumanMethylation 450K BeadChip (Illumina, Inc); EPIC BeadChip, Infinium Methylation EPIC BeadChip; WGS, whole genome sequencing. In MESA, participants who self-reported as Hispanic were analyzed alongside those reported as White, given that the genetic effects tend to be similar between these racial groups from a previous study.

**Supplemental Table 4**. Genomic control (λ_GC_) values in association analysis and meta-analysis of MSS and MH count

| **Study** | **Sample Size** | **Genomic Control Lambda** | |
| --- | --- | --- | --- |
|  |  | **MSS** | **MH count** |
| African Americans | | | |
| ARIC | 2114 | 1.08 | 0.77 |
| CARDIA | 989 | 1.00 | 1.01 |
| GENOA | 786 | 1.1 | 1.08 |
| JHS | 1706 | 1.03 | 0.92 |
| MESA | 174 | 1.46 | 1.38 |
| WHI EMPC | 113 | 1.29 | 1.59 |
| American Whites | | | |
| ARIC | 960 | 1.43 | 2.08 |
| CARDIA | 1268 | 1.08 | 1.05 |
| FHS | 1788 | 1.5 | 1.41 |
| MESA* | 680 | 1.12 | 0.84 |
| WHI BAA23 | 70 | 0.98 | 1.04 |
| WHI EMPC | 338 | 1.11 | 0.98 |
| Meta - analysis | | | |
| Pooled | 10,986 | 0.94 | 0.83 |
| EA | 5882 | 1.15 | 0.88 |
| AA | 5104 | 0.81 | 0.65 |

ARIC, Atherosclerosis Risk in Communities. CARDIA, Coronary Artery Risk Development in Young Adults. FHS, Framingham Heart Study. GENOA, Genetic Epidemiology Network of Arteriopathy. JHS, Jackson Heart Study. MESA, Multi-Ethnic Study of Atherosclerosis. WHI, Women’s Health Initiative

**Supplemental Table 5**. Comparison MHC and MSS scores: Pearson correlation coefficient

| cohort | Pearson correlation coefficient |
| --- | --- |
| FHS | 0.82 |
| CARDIA | 0.62 |
| MESA | 0.74 |
| JHS | 0.86 |
| WHI | 0.68 |
| GENOA | 0.89 |

ARIC, Atherosclerosis Risk in Communities. CARDIA, Coronary Artery Risk Development in Young Adults. FHS, Framingham Heart Study. GENOA, Genetic Epidemiology Network of Arteriopathy. JHS, Jackson Heart Study. MESA, Multi-Ethnic Study of Atherosclerosis. WHI, Women’s Health Initiative.

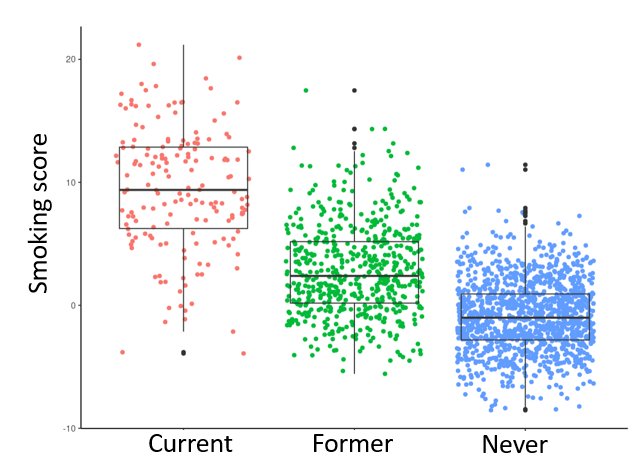

**Supplemental Figure 1**. The predicted smoking score vs. reported smoking status (current, former, and never). The smoking score was calculated from 183 CpGs using the EpiSmokEr R package.

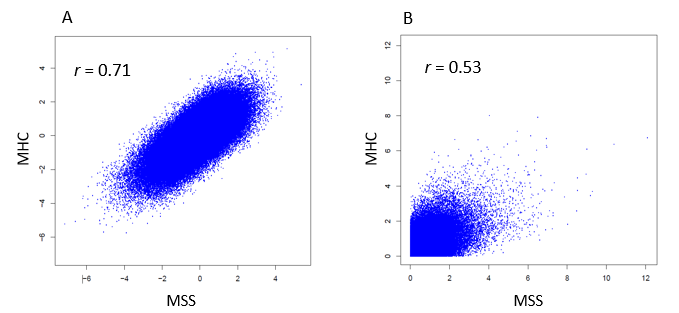

**Supplemental Figure 2**. Comparison of associations of DNA methylation with MSS and MHC. A. Comparison of effect magnitude (i.e., t value = beta/SE). B. Comparison of –log10 (p value).

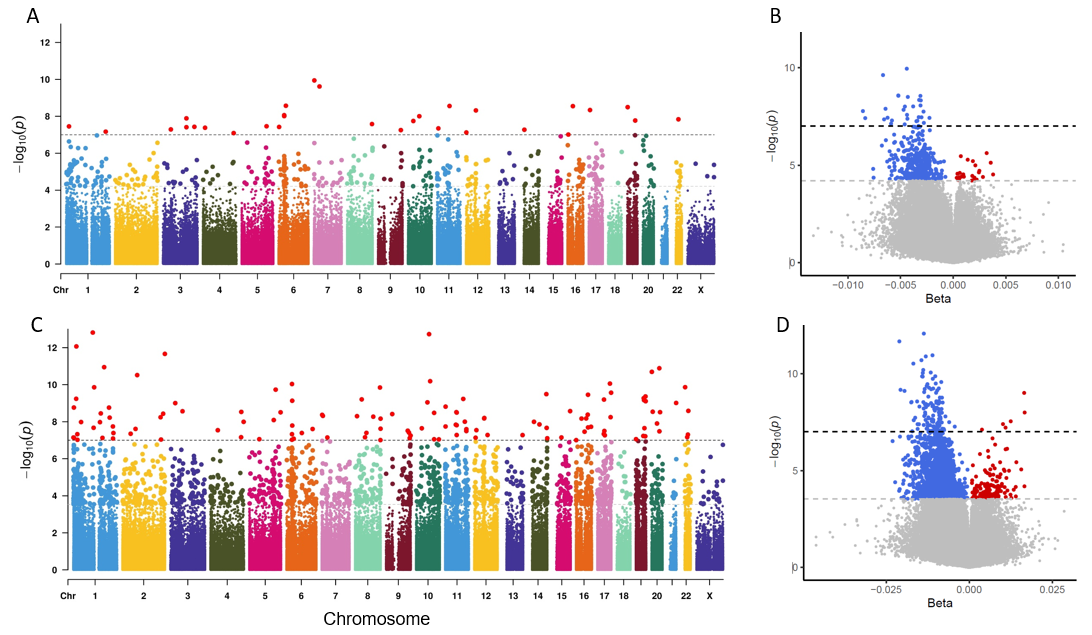

**Supplemental Figure 3**. Association and meta-analysis of American Whites (n=5104). A. MHC – Manhattan plot; B, MHC – volcano plot; C. MSS – Manhattan plot; D, MSS – volcano plot.

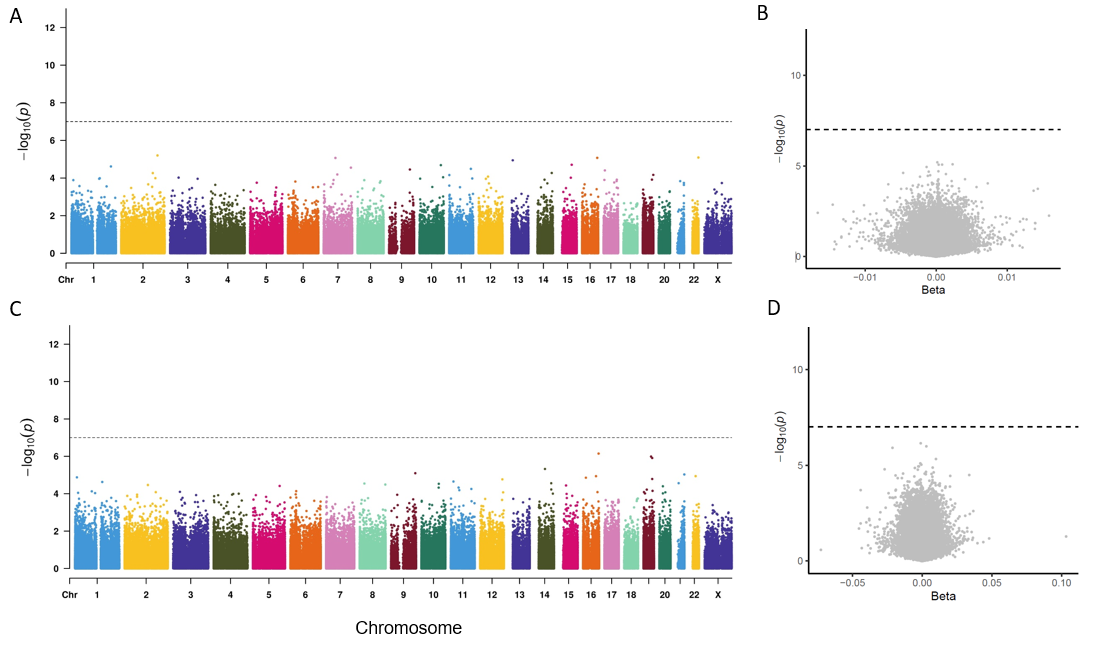

**Supplemental Figure 4**. Association and meta-analysis of participants of African American participants (n=5882). A. MHC – Manhattan plot; B, MHC – volcano plot; C. MSS – Manhattan plot; D, MSS – volcano plot

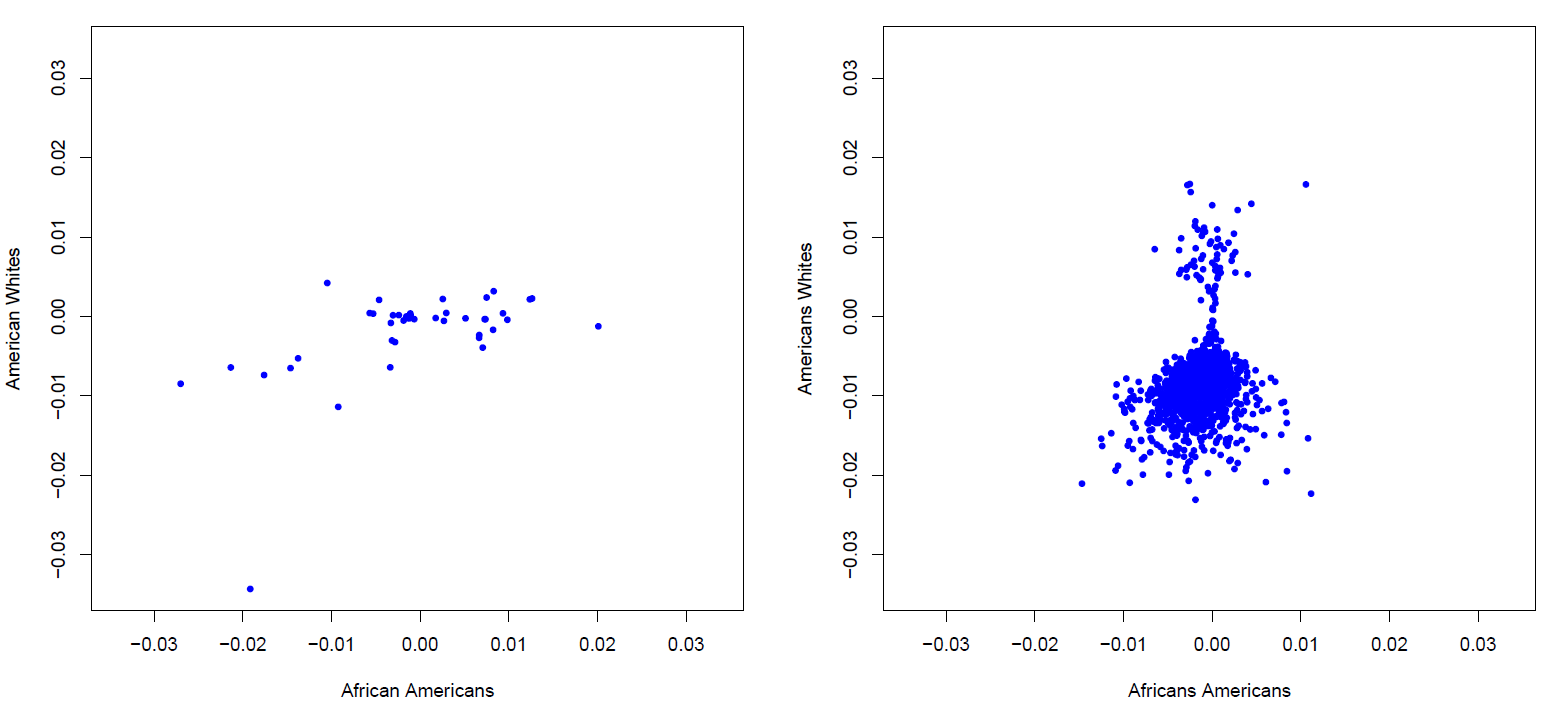

**Supplemental Figure 5**. Comparison of associations of DNA methylation with MSS between African Americans and American Whites. A. 44 CpGs selected in African Americans with p < 1e-4. B. 1295 CpGs in American Whites with p < 1e-4.

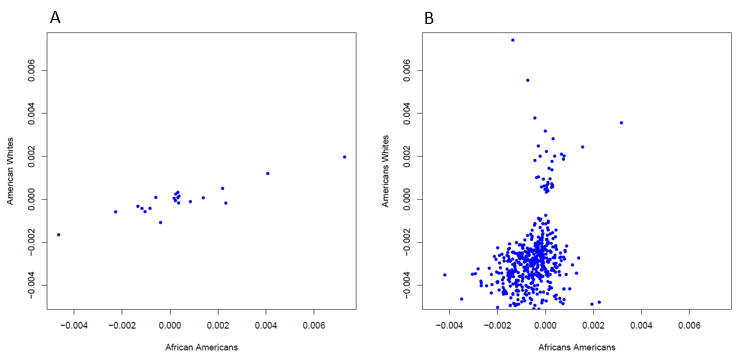

**Supplemental Figure 6**. Comparison of associations of DNA methylation with MHC between African Americans and American Whites. A. 21 CpGs selected in African Americans with p < 1e-4. B. 524 CpGs in American Whites with p < 1e-4.

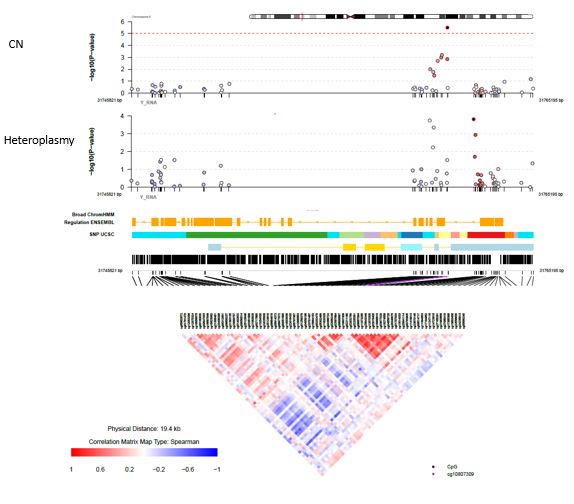

**Supplemental Figure 7**. Comparison between association analyses of DNA methylation with heteroplasmy analysis versus copy number in the VARS gene region on chromosome 6.

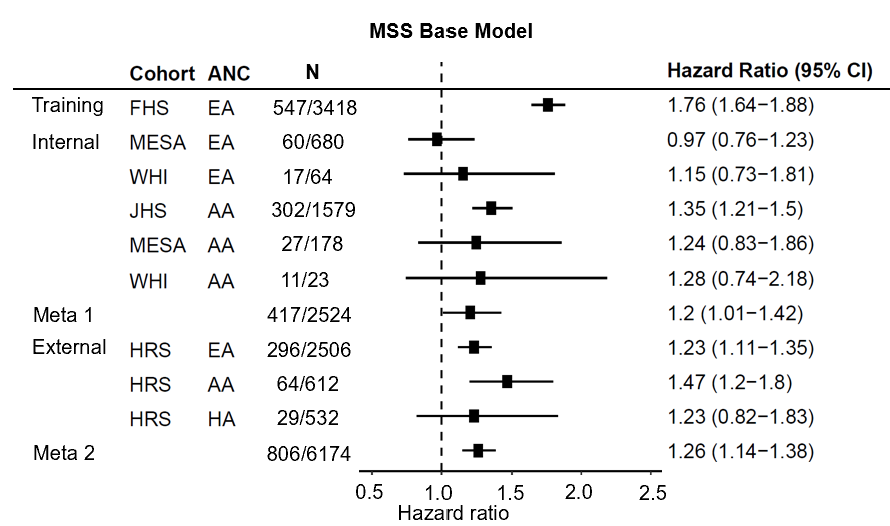

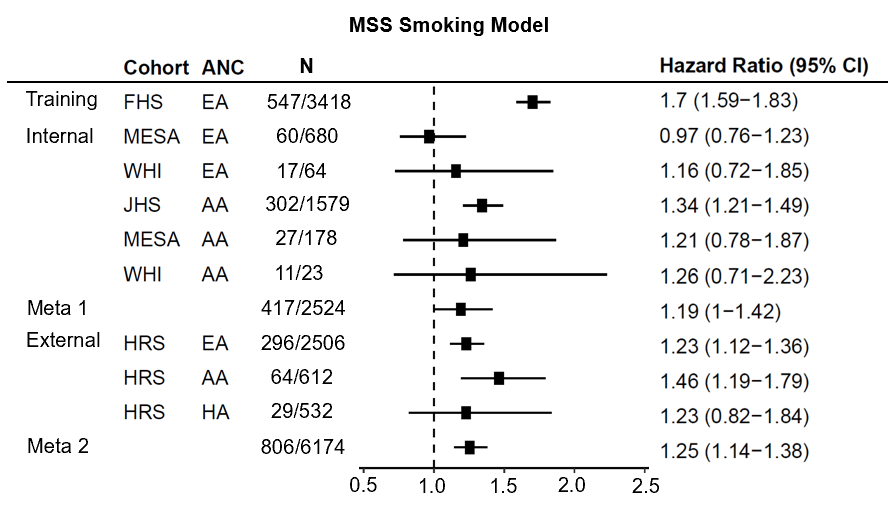

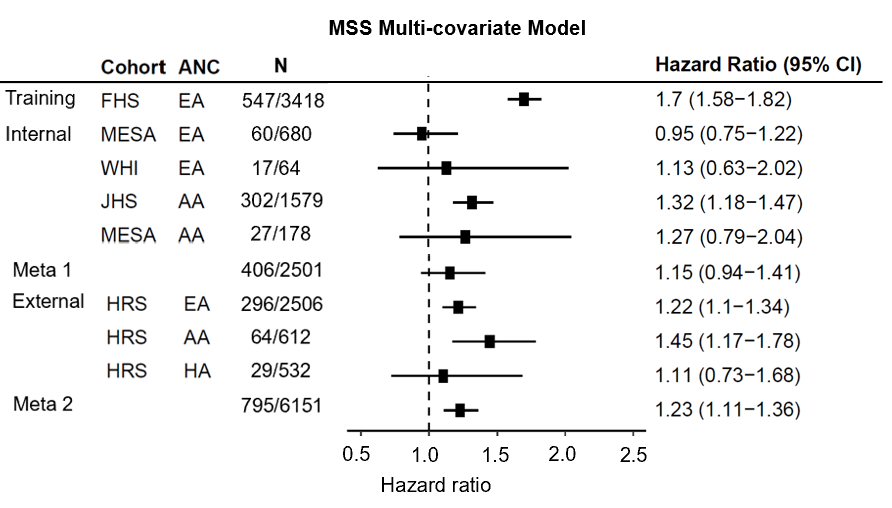

**Supplemental Figure 8**. **Forest plot: Association analysis of heteroplasmy-associated DNA methylation score (MSS) with all-cause mortality, adjusting for covariates.** We applied elastic net Cox regression to MSS associated CpGs and selected 56 CpGs for all-cause mortality. MSS, heteroplasmy burden score based on functional prediction of heteroplasmic variants. The forest plot illustrates hazard ratios (HR) with 95% confidence intervals (CI) for association analyses of the weighted MSS-CpG scores with mortality across the training and testing cohorts, adjusting for age and sex in the base model and multi-covariate model. In base model, covariates included age and sex. In smoking model, covariates included age, sex, and smoking status (never, former, and current). In multi-covariate model, covariates included age, sex, smoking status, BMI, systolic blood pressure, use of antihypertensive medication, total cholesterol, high-density cholesterol, diabetes, and use of lipid-lowering medication. Framingham Heart Study; JHS, Jackson Heart Study; HRS, Health and Retirement Study; MESA, Multi-Ethnic Study of Atherosclerosis Study, WHI, Women's Health Initiative.

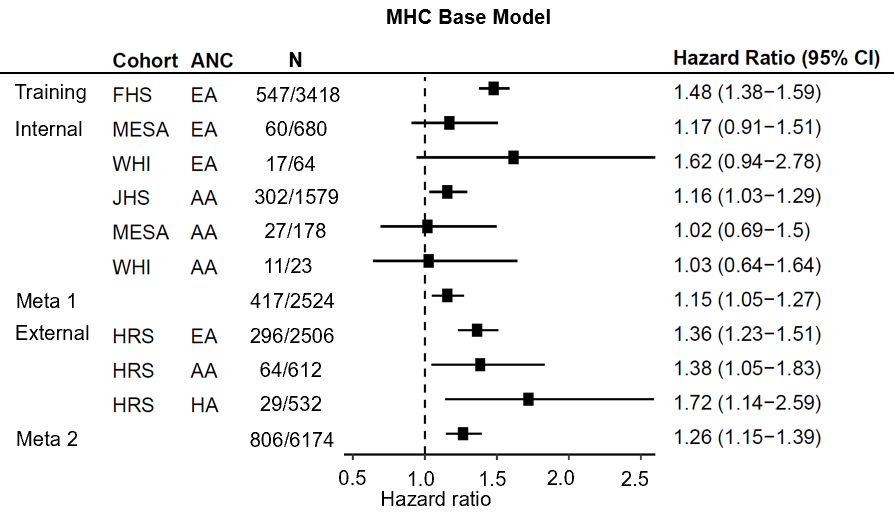

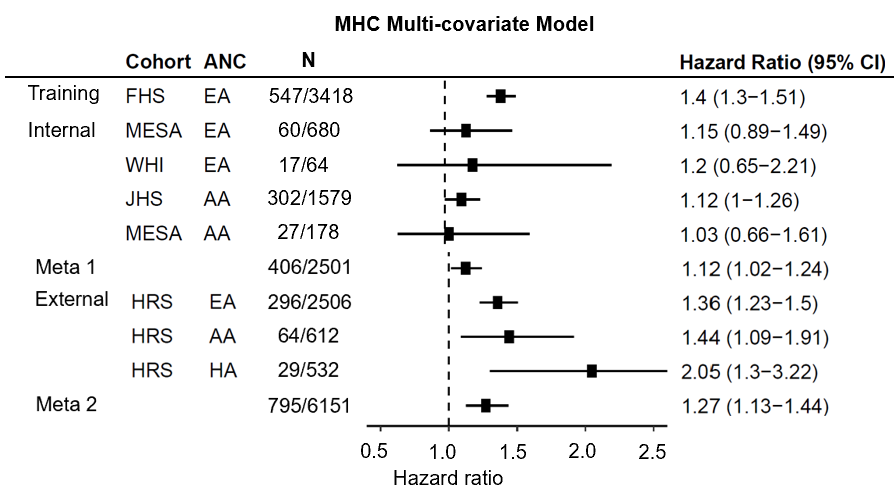

**Supplemental Figure 9**. **Forest plot: Association analysis of heteroplasmy-associated DNA methylation count score (MHC) with mortality, adjusting for covariates.** We applied elastic net Cox regression to MHC associated CpGs and selected 29 CpGs for all-cause mortality. The forest plot illustrates hazard ratios (HR) with 95% confidence intervals (CI) for association analyses of the weighted MHC-CpG scores with mortality across the training and testing cohorts, adjusting for age and sex in the base model and multi-covariate model. In base model, covariates included age and sex. In multi-covariate model, covariates included age, sex, smoking status, BMI, systolic blood pressure, use of antihypertensive medication, total cholesterol, high-density cholesterol, diabetes, and use of lipid-lowering medication. Framingham Heart Study; JHS, Jackson Heart Study; HRS, Health and Retirement Study; MESA, Multi-Ethnic Study of Atherosclerosis Study, WHI, Women's Health Initiative**.**

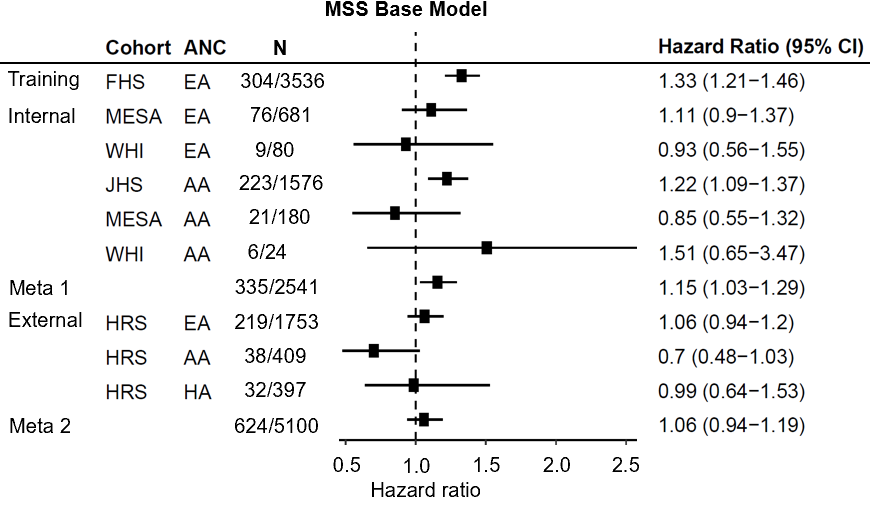

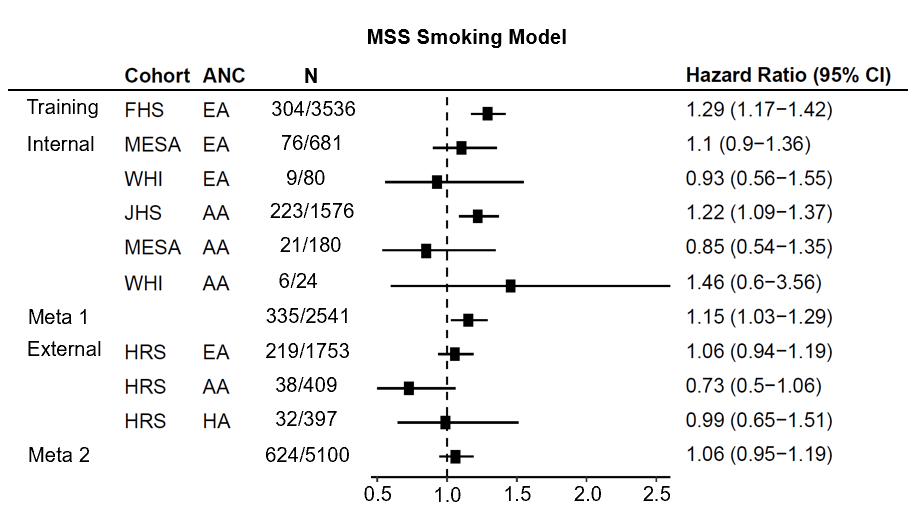

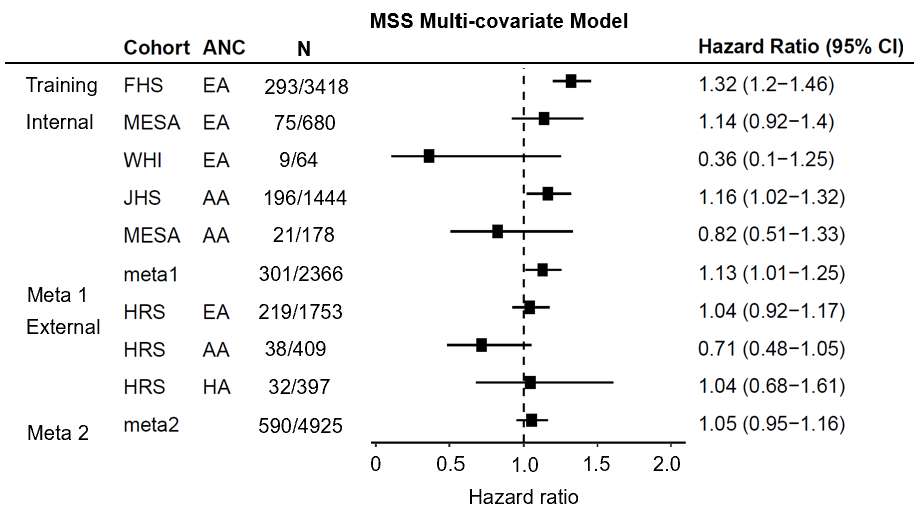

**Supplemental Figure 10**. **Forest plot: Association analysis of heteroplasmy-associated DNA methylation score (MSS) with CVD, adjusting for covariates.** We applied elastic net Cox regression to MSS associated CpGs and selected 9 CpGs for CVD. MSS, heteroplasmy burden score based on functional prediction of heteroplasmic variants. The forest plot illustrates hazard ratios (HR) with 95% confidence intervals (CI) for association analyses of the weighted MSS-CpG scores with mortality across the training and testing cohorts, adjusting for age and sex in the base model and multi-covariate model. In base model, covariates included age and sex. In smoking model, covariates included age, sex, and smoking status (never, former, and current). In multi-covariate model, covariates included age, sex, smoking status, BMI, systolic blood pressure, use of antihypertensive medication, total cholesterol, high-density cholesterol, diabetes, and use of lipid-lowering medication. Framingham Heart Study; JHS, Jackson Heart Study; HRS, Health and Retirement Study; MESA, Multi-Ethnic Study of Atherosclerosis Study, WHI, Women's Health Initiative.

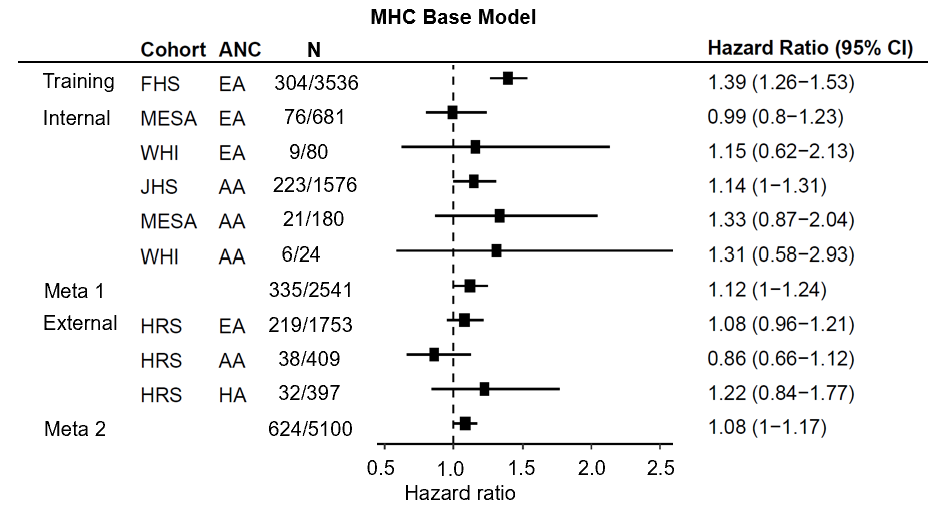

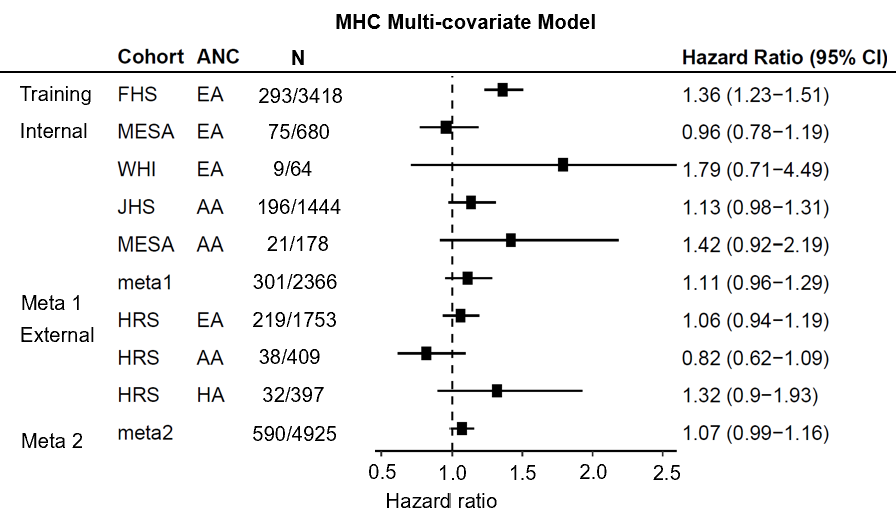

**Supplemental Figure 11**. **Forest plot: Association analysis of heteroplasmy-associated DNA methylation score (MHC) with CVD, adjusting for covariates.** We applied elastic net Cox regression to MHC associated CpGs and selected 13 CpGs for all-cause mortality. The forest plot illustrates hazard ratios (HR) with 95% confidence intervals (CI) for association analyses of the weighted MHC-CpG scores with CVD across the training and testing cohorts, adjusting for age and sex in the base model and multi-covariate model. In base model, covariates included age and sex. In multi-covariate model, covariates included age, sex, smoking status, BMI, systolic blood pressure, use of antihypertensive medication, total cholesterol, high-density cholesterol, diabetes, and use of lipid-lowering medication. Framingham Heart Study; JHS, Jackson Heart Study; HRS, Health and Retirement Study; MESA, Multi-Ethnic Study of Atherosclerosis Study, WHI, Women's Health Initiative.
